# Diverse mediators of cancer predisposition uncovered by germline whole genome sequencing of unexplained familial cancers

**DOI:** 10.64898/2026.05.08.26352653

**Authors:** Noah Fields, Seunghun Han, Wenbin Mei, Erin Shannon, Ryan Buehler, Deborah Neklason, Erica Pimenta, Junne Kamihara, Judy Garber, Riaz Gillani, Saud AlDubayan, Jihye Park, Ryan L. Collins, Eliezer M. Van Allen

**Affiliations:** Department of Medical Oncology, Dana-Farber Cancer Institute, Boston, MA, USA; Cancer Program, Broad Institute of Harvard and MIT, Cambridge, MA, USA; Department of Biological and Biomedical Sciences, Harvard Medical School, Boston, MA, USA; Division of Cancer Genetics and Prevention, Dana-Farber Cancer Institute, Boston, MA, USA; Utah Center of Genetic Discovery, University of Utah, Salt Lake City, UT, USA; Huntsman Cancer Institute, University of Utah, Salt Lake City, UT, USA; Harvard Medical School, Boston, MA, USA; Department of Pediatric Oncology, Dana-Farber Cancer Institute, Boston, MA, USA; Division of Hematology/Oncology, Boston Children’s Hospital, Boston, MA, USA; Division of Genetics, Brigham and Women’s Hospital, Boston, MA, USA; College of Medicine, King Saud bin Abdulaziz University for Health Sciences, Riyadh, Saudi Arabia

## Abstract

Cancer frequently clusters in families due to shared environment and genetics. However, many familial cancer cases lack a clinically recognized pathogenic germline variant (PGV). We analyzed germline genomes and family history from 2,726 individuals without a PGV in the All of Us Research Program, including 1,496 cases across 18 cancer types with extensive family history and 1,230 family history-negative, cancer-free controls. We identified allelic series of rare structural variants inactivating *MSH2* in individuals with phenotypes consistent with Lynch syndrome and *BRCA1* in breast cancer. Cancer polygenic risk scores were enriched in cases and correlated with patterns of cancer diagnoses within families. Exome-wide rare variant analyses nominated six candidate predisposition genes, including *TSTD2* and *BRAT1* in thyroid and breast cancer, respectively. Overall, polygenic risk and rare variants impacting known genes explained a median of 5% of unexplained familial cancers, increasing to 11% when including newly nominated risk factors.

## INTRODUCTION

While environmental carcinogens and lifestyle factors contribute to the development of cancer, heritable genetic factors are also a major source of cancer risk^1,2^. For example, a seminal study from The Cancer Genome Atlas (TCGA) reported that approximately 8% of unselected adult cancer patients carry a rare pathogenic germline variant (PGV) in a known cancer predisposition gene (CPG)^3^. The TCGA study reported PGVs in 9.9% of breast cancers; however, this prevalence rises to nearly 30% when restricting to familial breast cancers^4^. Although pan-cancer studies of PGVs in high-risk families are limited, similar trends have been observed in colorectal cancer, with ~25% of familial cases carrying a PGV^5^. Together, these results suggest that (i) a family history of cancer strongly enriches for genetic predisposition, and (ii) many families with extensive cancer histories still lack an identifiable genetic cause. This highlights our current gap in understanding of the inherited causes of cancer that likely lie beyond conventional PGVs impacting established CPGs.

Standard clinical germline cancer risk assessment for PGVs typically involves targeted sequencing of a panel of known CPGs and subsequent analysis of primarily coding single nucleotide variants (SNVs) and small insertions/deletions (indels), with a minority of panels reporting some large deletions and duplications. These targeted approaches fail to capture the majority of variation present in a patient’s germline genome, including structural variants (SVs; rearrangements ≥50bp) and variants in the ~98% of the human genome that do not encode proteins but may affect gene regulation and splicing^6,7^. Furthermore, many clinical genetic assessments do not incorporate polygenic risk, which measures the cumulative effect of many common low-risk variants^8,9^. Germline whole-genome sequencing (WGS) uniquely addresses these limitations by comprehensively profiling a patient’s entire inherited genome to simultaneously detect variants across contexts (coding & noncoding), frequencies (common & rare), and mutational classes (SNVs, indels, and SVs)^10,11^. Germline WGS may therefore provide a more comprehensive view of an individual’s inherited cancer susceptibility, which will be especially necessary for unexplained familial cancers^12^.

The emergence of nation-scale research biobanks, like the All of Us Research Program (All of Us), now provide large, diverse cohorts with germline WGS paired to electronic health records (EHR), survey responses, and other phenotypic data^13,14^. In this study, we leveraged All of Us to retrospectively identify 1,496 individuals diagnosed with at least one primary cancer who also (1) reported significant family history of cancer and (2) lacked an established SNV/indel PGV in a known CPG. We defined a broad spectrum of germline variation in these individuals and a covariate-matched set of 1,230 cancer-free, PGV-negative controls with no reported cancer family history to quantify familial cancer risk contributed by multiple historically understudied mechanisms, including rare SVs, polygenic risk, cryptic splice variants, variants of uncertain significance, and runs of homozygosity. Collectively, our analyses suggest familial cancers arise from a wider spectrum of both common and rare germline variation than has been previously appreciated, with immediate implications for translational studies and clinical management of high-risk families.

## RESULTS

### Generation and characterization of an unexplained familial cancer cohort from the All of Us Research Program

We integrated short-read germline WGS, EHR, and survey data available for 203,354 individuals in the All of Us Research Program Controlled Tier v7 (CDR v7) to retrospectively define a cohort of 1,496 unexplained familial cancer cases and 1,230 cancer-free, family history-negative controls (**Fig. 1a**). We defined candidate cases as individuals with a documented cancer diagnosis and at least three affected first- or second-degree relatives, while candidate controls had no known cancer diagnosis and no reported cancer in first- or second-degree relatives. We subsequently excluded all individuals carrying a PGV or rare loss-of-function (LoF) variant in any of 148 CPGs^15^ (**Fig. 1b**; **Table S1**), reasoning that this criterion roughly approximates current clinical germline testing for suspected hereditary cancer. These filters retained 1,496 cases, which we further grouped based on EHR diagnostic codes into 18 distinct cancer types (**Table S2**), and in parallel generated covariate-matched controls for each cancer type by subsetting from our 1,230 eligible controls. Most cases (66%) mapped to just one cancer, 26% of cases mapped to multiple cancers even after accounting for common metastatic sites, and the remaining 8% had either ambiguous EHR encodings or rarer cancers not analyzed here. Notably, we did not enforce any requirement for each case’s cancer diagnosis to match their cancer family history, yet our ascertainment criteria enriched for families with specific cancers relative to the general population, likely reflecting a combination of shared genetic susceptibility, environmental exposures, and lifestyle factors (**Fig. 1c**; **Fig. S1**). To further quantify the patterns of familial clustering of specific cancers in this cohort, we tested for associations between first-degree relative (FDR) reported diagnosis of 14 different cancers and case diagnosis of 18 different cancers using logistic regression adjusted for sex. The 11 strongest associations all involved enrichments (odds ratios [ORs] > 1) of matched cancers between patients and their FDRs, which may reflect underlying genetic predisposition even in the absence of PGVs (**Fig. 1d,e**).

**Figure 1.**
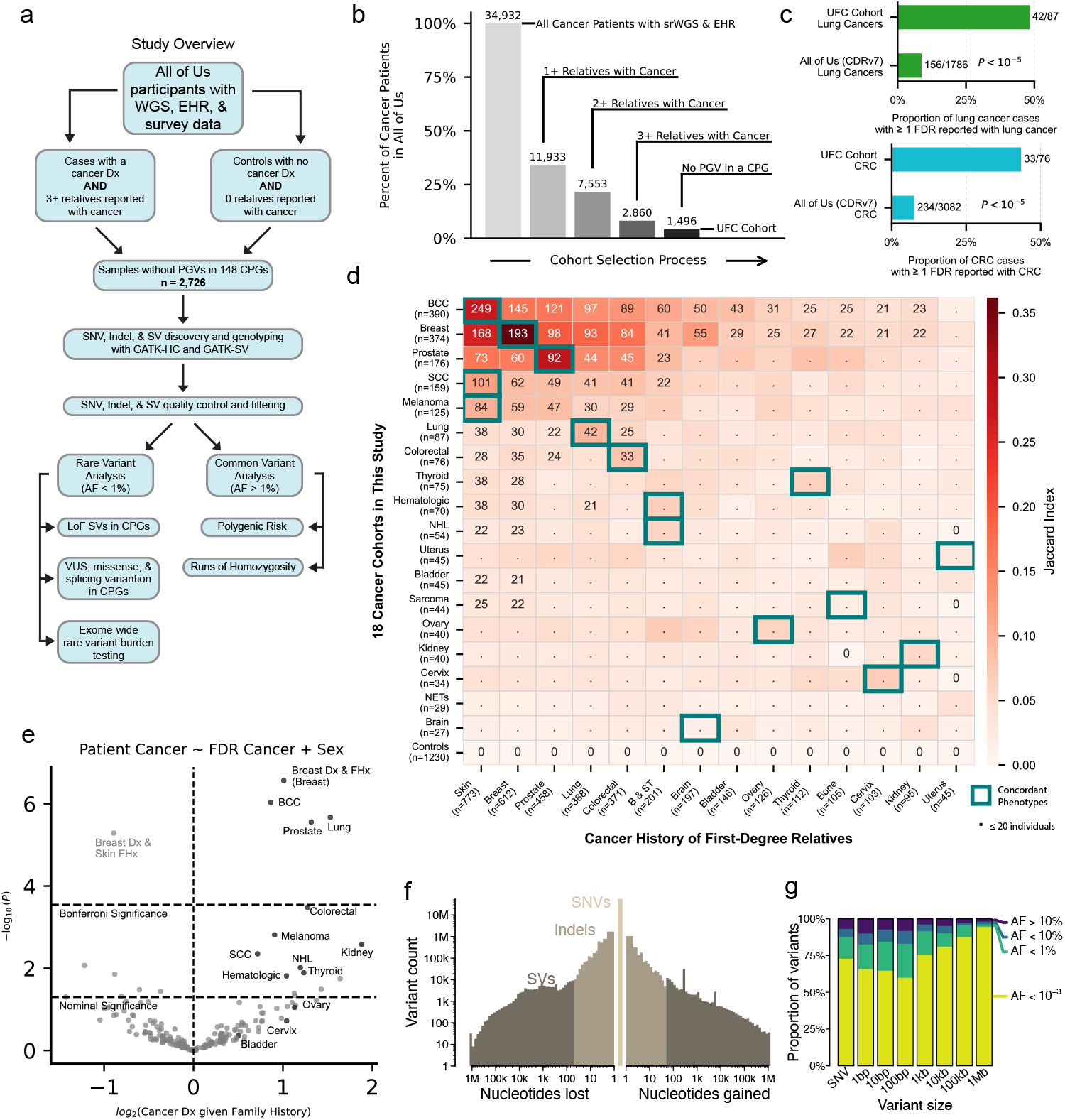
Retrospective construction of an unexplained familial cancer cohort from the All of Us Research Program. **(a)** Roadmap of cohort creation, quality control, and primary analyses. **(b)** Progressive filtering criteria applied to All of Us CDR v7 to identify 1,496 unexplained familial cancer (UFC) cases lacking a PGV. srWGS., short read whole-genome sequencing. **(c)** Lung and colorectal cancer (CRC) cases in the UFC cohort show higher rates of concordant family history compared to the broader All of Us cohort. **(d)** Correspondence of patient cancer diagnosis and self-reported family history. BCC., basal cell carcinoma; SCC., squamous cell carcinoma; NHL., non-Hodgkin lymphoma; NETs., neuroendocrine tumors; B & ST., blood and soft tissue tumors. **(e)** Association testing of all 18 cancer types categorized in patients versus 14 cancer types reported in first-degree relatives (FDRs). **(f)** Our germline WGS pipelines captured the full spectrum of variant sizes, spanning from SNVs to large SVs. **(g)** Most germline variants in this cohort were rare, with AF of large SVs correlating with size as expected.

We next performed genome-wide discovery and genotyping of SNVs, indels, and SVs using GATK-HC^16^ and GATK-SV^17^. These methods collectively captured 89 million total variants present in at least one genome after rigorous quality control and filtering (**Fig. 1f, Fig. S2, S3**), which translated to an average of 3.6 million SNVs, 394 thousand indels, and 9,257 SVs per genome. Consistent with prior population sequencing studies, 87% of the SNVs identified in this cohort were rare (allele frequency [AF] < 1%) (**Fig. 1g**). We assessed ancestral population structure using principal component analysis, finding that 85% of individuals most closely matched European genetic ancestry groups (**Fig. S4, S5**).

### Rare SVs impact known CPGs and are enriched in a subset of unexplained familial cancers

Germline SVs are a major source of gene-disruptive variation, accounting for at least one-quarter of all genic LoF events per genome^17,18^. To assess the contribution of SVs at WGS resolution in unexplained familial cancers, we comprehensively searched the 1,489 cases and 1,206 controls (99% of cohort) passing strict SV-specific quality control for SVs predicted to result in LoF of a known CPG. We identified 45 ultra-rare (AF < 0.1%) SVs predicted to result in LoF of 33 different CPGs. These ultra-rare LoF SVs were identified in 55 individuals (2.04% of all samples; 95% confidence interval [CI] = 1.54–2.65%) (**Fig. 2a-e**). Eight CPGs were impacted by two or more distinct ultra-rare LoF SVs, including three independent LoF SVs in both *MSH2* and *BRCA1*. Notably, the *MSH2* SVs were identified in patients with colorectal, ovarian, and uterine cancers, suggesting that these SVs may represent an underlying genetic cause in individuals with Lynch-like phenotypes^19^. Due to regulatory requirements, all research studies using All of Us data, including ours, are prohibited from reporting participant counts ≤20; hence, we could not report cancer type-specific rates in this study. Nonetheless, we calculated the aggregate burden of rare LoF SVs affecting CPGs for each of the 18 cancer types in our cohort, stratifying variants by increasingly strict AF thresholds. The prevalence of rare LoF SVs impacting known CPGs differed across cancer types. Although rare SVs were 1–2 orders of magnitude sparser than SNVs/indels, we nonetheless identified nominally significant excesses of singleton LoF SVs in CPGs for four cancer types (P < 0.05; Fisher’s exact test). Across nearly all cancer types, we consistently observed increased enrichments of rare LoF SVs in cases versus controls that inversely correlated with stricter AF thresholds (*e*.*g*., in neuroendocrine tumors: OR=2.6 for SVs at AF <1% vs. OR=9.0 for singleton SVs). These results have implications for the interpretation of germline SVs in clinical germline testing for hereditary cancers, where lower population AFs imply a higher likelihood of pathogenicity.

**Figure 2.**
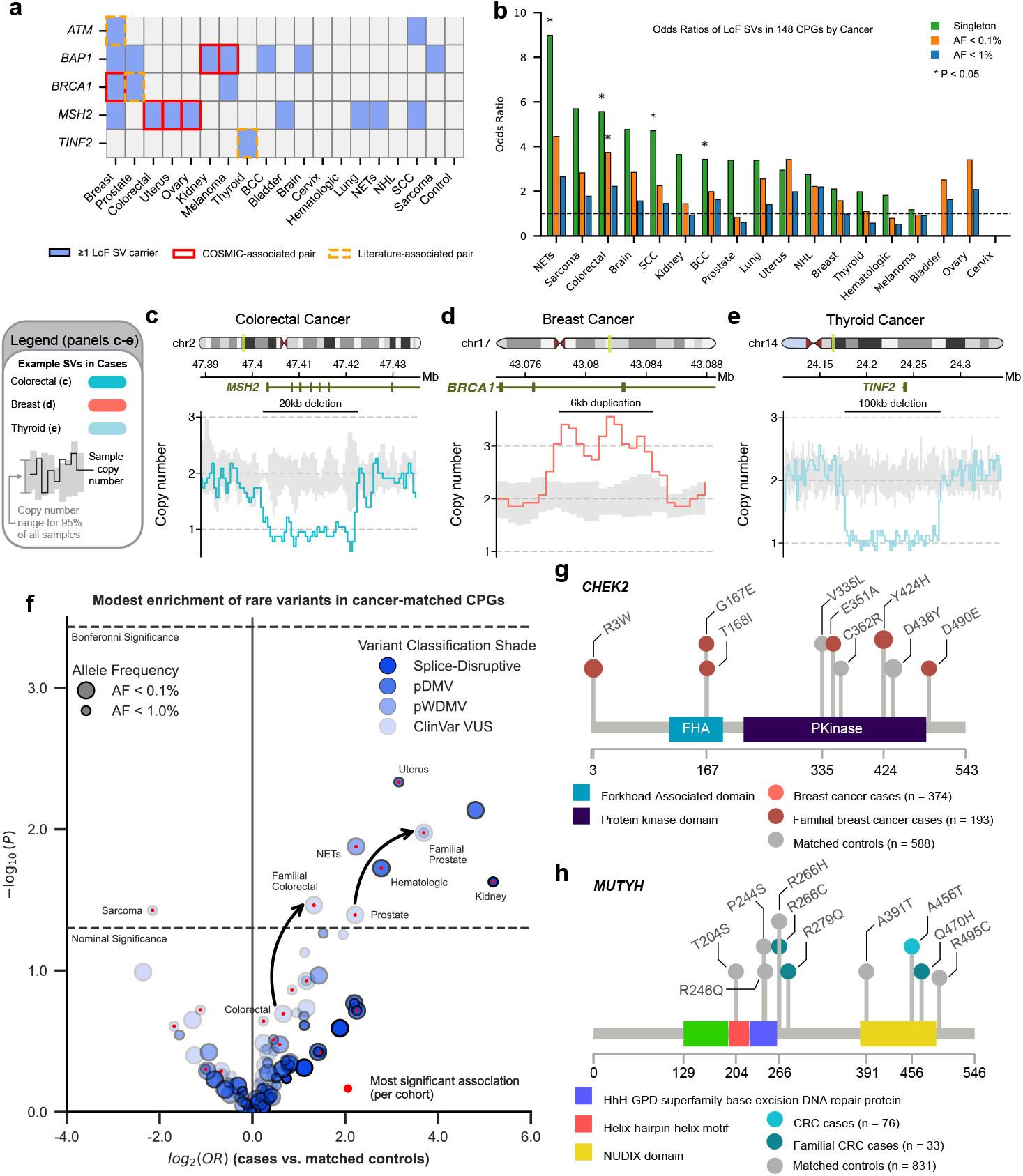
Unexplained familial cancers harbor LoF SVs and rare SNVs/ indels of uncertain significance in CPGs. **(a)** Matrix of rare (AF < 1%) LoF SVs across five selected CPGs in 18 cancer types and controls. Blue rectangles highlight that at least one individual with a LoF SV in that gene has been diagnosed with the cancer type on the x-axis. A COSMIC-associated pair denotes a gene–cancer germline predisposition association curated by the Catalogue of Somatic Mutations in Cancer (COSMIC)^73^, whereas a literature-associated pair denotes an association not represented in COSMIC but described in the published literature^30,74^. **(b)** Effect sizes of rare LoF SVs in unexplained familial cancer cases relative to matched controls were inversely correlated with AF, with rarer LoF SVs exhibiting greater enrichments in cases. Example LoF SVs included **(c)** a 20kb deletion in MSH2 in a colorectal cancer case, **(d)** a 6kb duplication in BRCA1 in a breast cancer case, and **(e)** a 100kb deletion encompassing TINF2 in a thyroid cancer case. **(f)** Rare coding and splicing variants in CPGs were modestly enriched across multiple cancer types at nominal significance. Two classifications for missense variants were analyzed: pDMV., predicted damaging missense variant; pWDMV., predicted weakly damaging missense variant. Gene-centric analyses revealed focused enrichments of ultra-rare (AF < 0.1%) missense variants in **(g)** CHEK2 in breast cancer and **(h)** MUTYH in colorectal cancer (CRC). Protein residues corresponding to observed missense variants are colored by family history status, with darker shades indicating variants identified in at least one case with concordant family history (all rare CHEK2 missense variants tracked with familial breast cancer). Control-only variants were shown in gray, and no variant was shared between cases and controls.

### Modest excesses of rare variants of uncertain clinical significance in known CPGs

Interpreting rare variants in CPGs is challenging when their functional impact is unclear and they are either absent from clinical databases like ClinVar^20^ or present in ClinVar but annotated as “variants of uncertain significance” (VUSs). We hypothesized that a subset of currently uninterpretable rare variants in known CPGs are pathogenic, and that rare VUSs might therefore be enriched in CPGs in cases in our cohort that was pre-screened for established PGV SNV/indels. We thus evaluated (i) rare variants annotated as VUSs in ClinVar, (ii-iii) rare missense variants in CPGs predicted to be (ii) damaging or (iii) weakly damaging by *in silico* algorithms (i.e., AlphaMissense^21^, REVEL^22^, or PrimateAI^23^), and (iv) rare splice-disruptive variants predicted to be pathogenic by SpliceAI^24^. We further refined our CPG list for these targeted analyses by matching against COSMIC’s (Catalogue of Somatic Mutations in Cancer)^25^ cancer gene census to obtain 139 curated gene-cancer pairs across 79 genes (**Table S3**). Cervical cancer had no CPGs listed in COSMIC and therefore was excluded from this analysis.

We tested for associations between the four rare variant categories defined above in cancer type-matched CPGs at two frequency thresholds (AF < 1% and 0.1%) and case–control status for 17 cancer types. Given our relatively small sample sizes and CPG lists for most cancer types, no comparison surpassed strict Bonferroni-adjusted significance (P < 2.9 × 10^−3^). However, 5/17 cancer types exhibited nominally significant enrichments of rare variants in CPGs in at least one category (**Fig. 2f**; P < 0.05; logistic regression adjusted for sex and ancestry). Moreover, as recent studies have reported increased rates of rare PGVs in patients with concordant family histories^4,5^, we further hypothesized that restricting these analyses to cases with matched family history would enhance signals of true pathogenicity in CPGs. Although our sample sizes were limited, we defined subsets of cases with concordant family history for four cancer types: colorectal (n = 33) and lung (n = 42) cases with ≥1 affected FDR, and breast (n = 41) and prostate (n = 27) cases with ≥2 affected FDRs. Enforcing concordant family history and patient cancer diagnoses markedly strengthened the association between prostate cancer and ClinVar VUSs in CPGs (OR = 12.93; P = 1.06 × 10^−2^) and yielded a newly nominally significant association between colorectal cancer and rare ClinVar VUSs in CPGs (OR = 2.51; P = 3.45 × 10^−2^). One limitation of gene set-based analyses is that gene-specific effects may be obscured within large gene sets because signals are aggregated across genes with heterogeneous effect sizes and variant burdens. To address this, we examined individual genes and observed suggestive enrichment patterns. For example, we identified six ultra-rare (AF < 0.1%) missense variants in *CHEK2* in breast cancer patients, and four such variants in *MUTYH* in colorectal cancer patients (**Fig. 2g, h**). Collectively, these results suggest that a small subset of unexplained familial cancer cases harbor rare variants in CPGs that are currently clinically undefined but may be pathogenic.

### Common polygenic risk for cancer is enriched in unexplained familial cancer patients and tracks with specific family histories

Familial cancer studies have traditionally focused on rare, highly penetrant variants that follow Mendelian segregation patterns within pedigrees. However, increasingly large genome-wide association studies of cancer cases and controls have demonstrated a major risk contribution from the aggregated effects of common risk variants littered throughout the genome, usually summarized as a polygenic risk score (PRS). We therefore hypothesized that some unexplained familial cancers may reflect families that carry exceptionally high polygenic risk. We tested this hypothesis by selecting four published studies that uniformly developed 39 PRS models matching 14 of our 18 cancer types^26–29^. We applied each PRS model to its corresponding cancer type in our cohort, adjusted scores for the top 4 generrtic principal components to account for differences driven by ancestry, and tested case–control associations using logistic regression. In 8 of 14 cancer types, at least one PRS model was nominally associated with case–control status (P < 0.05), with 5 remaining significant after strict Bonferroni correction (P < 4 × 10^−3^; **Fig. 3a**). Odds ratios for the 8 significant PRS ranged from 1.4 to 2.2 per standard deviation increase in PRS (median = 1.5). The lack of significant associations between matched PRS and 6 cancer types likely reflects limited sample sizes, phenotypic mismatches (*e*.*g*., glioblastoma PRS applied to all brain cancers), and strict cohort ascertainment; however, all six exhibited OR > 1 with their best phenotype-matched PRS. Given potential confounding of PRS by ancestry and population structure, we repeated these analyses after restricting to individuals of European genetic ancestry, finding no meaningful differences in any trends. Although we evaluated all 39 PRS models within their respective cancer types, subsequent analyses were restricted to the most significant model per cancer type (lowest p-value).

**Figure 3.**
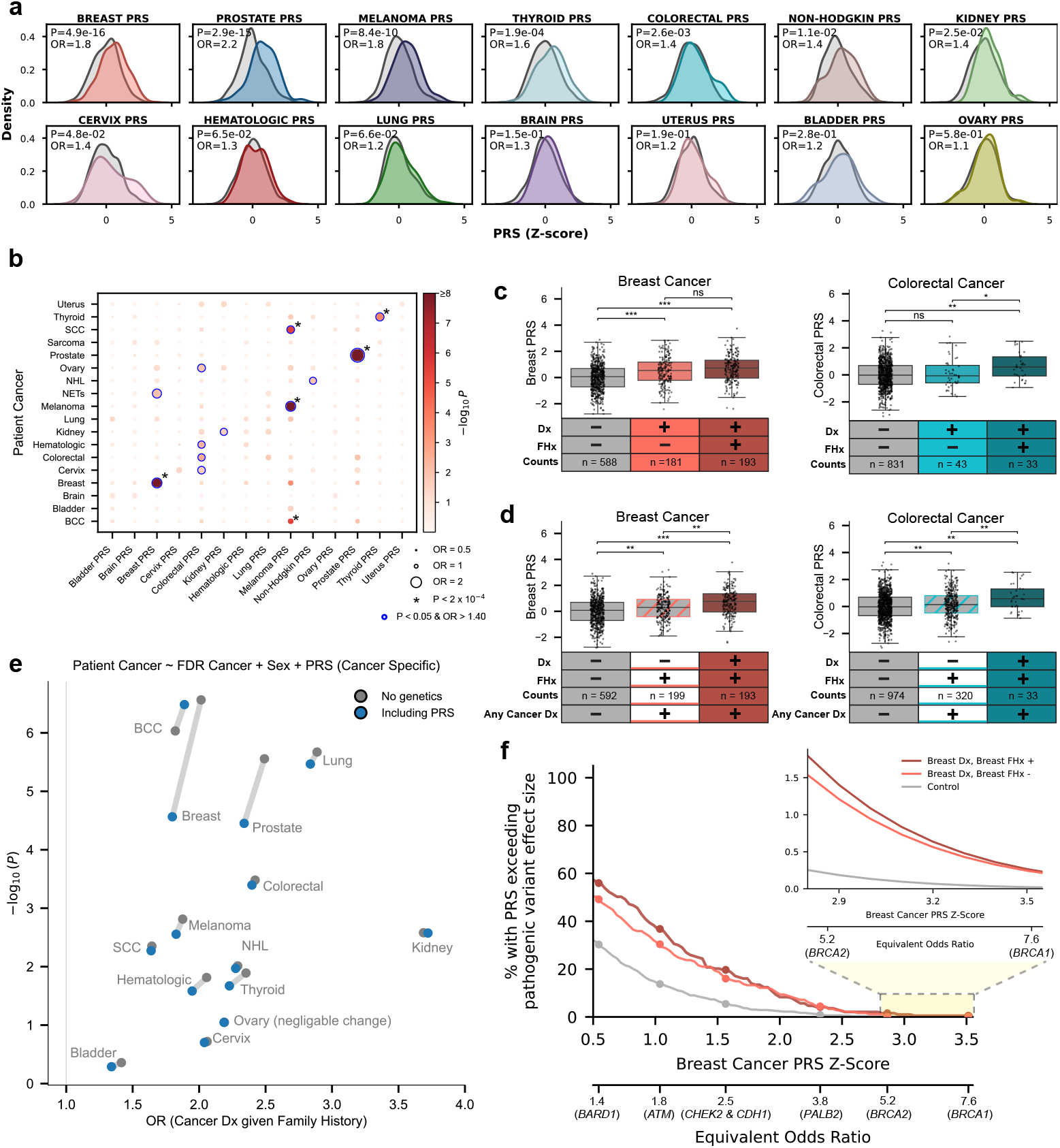
Common variant polygenic risk for cancer is concentrated in families with unexplained cancer histories. **(a)** The distribution of cancer type-matched PRS across 14 cancers in this study. Controls are represented in gray. **(b)** Summary of complete association testing of all 14 cancer type-specific PRS models versus all 18 cancer types in this study. BCC., basal cell carcinoma; NETs., neuroendocrine tumors; NHL., non-Hodgkin lymphoma; SCC,. squamous cell carcinoma. * indicates Bonferroni-adjusted significance (P < 2 × 10^−4^), correcting for 18 cancers and 14 PRS models (252 tests). **(c)** Polygenic risk is elevated in cases with concordant family history (FHx) in first-degree relatives. Statistical significance is indicated as follows: P < 0.05 (*), P < 0.01 (**), P < 1 × 10^−5^ (***), and not significant (ns). **(d)** Cases diagnosed with the PRS-matched cancer inherited elevated PRS compared to individuals with comparable family history who developed other cancers. **(e)** PRS partially attenuated the association between an individual’s cancer diagnosis and their FDR’s cancer diagnosis. **(f)** A small fraction of breast cancer cases displayed extremely elevated PRS, conferring risk comparable to PGVs in highly penetrant breast cancer CPGs. The smoothed curve in the upper right represents the estimated proportion of individuals with PRS above a given threshold, derived from the normal distribution of PRS. To adhere to the All of Us data and statistics dissemination policy, we have omitted exact counts and percentages from the upper right sub-panel.

Because many hereditary cancer syndromes predispose to cancers across multiple organs, we next explored whether the PRS for each cancer was associated with case–control status across the other 17 cancer types evaluated in our study. Each of the eight PRS models that were nominally significant when applied to its intended cancer type showed its strongest association with that cancer (**Fig. 3b**). Exceptional specificity was observed in prostate, thyroid, kidney, and non-Hodgkin lymphoma PRSs, which were exclusively associated with case–control status in their matched cancers (P < 0.05 and OR > 1.40; **Table S4, S5**).

Building on the apparent specificity of PRS models, we next evaluated whether PRS differed between cases stratified by concordant vs. discordant cancer family histories among FDRs. In 9 of the 12 cancer types with sufficient sample sizes, mean PRS was higher in cases with concordant vs. discordant family history, a trend that was nominally significant in colorectal (P = 1.17 × 10^−2^), kidney (P = 2.23 × 10^−3^), and hematologic (P = 1.34 × 10^−2^) cancers (one-sided Student’s t-test; **Fig. 3c**; **Fig. S6**). We next tested the reciprocal hypothesis: whether PRS differed among individuals with comparable cancer family history but divergent personal cancer diagnoses. For example, among individuals reporting a FDR with colorectal cancer, colorectal cancer PRS was nominally higher in colorectal cancer cases than in those who developed other cancer types (P = 3.13 × 10^−3^; one-sided Student’s t-test). This pattern was consistent across analyses: in 8 of 9 eligible cancer types, among individuals with comparable cancer family history, the mean PRS was higher in those whose cancer matched that of their affected FDRs than in those with discordant cancer diagnoses. (**Fig. 3d; Fig. S7**). Given this strong relationship between family history and PRS, we asked to what extent our previously observed clustering of specific cancers within families could be explained by PRS. We incorporated cancer-specific PRS into logistic regression models across the same 14 cancer types, finding that PRS attenuated the association between family history and patient diagnosis for 12 of 14 cancer types (**Fig. 3e**). To assess the clinical utility of these PRS results, we calculated the proportion of breast cancer cases with a breast cancer PRS exceeding the risk conferred by PGVs in CPGs implicated in breast cancer in the general population^30^. In our cohort, PRS conferred a higher risk than a *CHEK2* PGV in 19.7% of familial breast cancer cases and a higher risk than a *BRCA1* PGV in a small subset of these cases (**Fig. 3f**). Collectively, these results indicate a prominent and specific role for common variants in familial aggregation in otherwise “unexplained” cancers.

### Systematic discovery of new candidate cancer predisposition genes

In addition to the features already explored, another potential explanation for unexplained familial cancers is the presence of rare pathogenic coding and splicing variants in as-of-yet undiscovered CPGs. To search for new CPGs, we first classified all rare genic SNVs/indels into four tiers of predicted severity using a combination of five *in silico* methods plus ClinVar annotations: Tier 1, high-confidence loss-of-function (LoF); Tier 2, low-confidence LoF and predicted splice-disruptive variants; and Tiers 3–4, missense variants stratified by predicted pathogenicity (**Fig. S8**). We then performed collapsing rare variant burden tests at two different frequency thresholds (AF < 1% and 0.1%) for each of Tiers 1–4 across all 18,544 autosomal protein-coding genes. Because rare variant analyses are sensitive to small case counts, we restricted these analyses to 15/18 cancer types with at least 40 cases. Using SAIGE-GENE+^31^, we assessed rare variant burden in each of 15 cancers by comparing cases to controls while adjusting for sex and ancestry.

In total, our exome-wide discovery analyses nominated 14 candidate genes associated with at least one cancer at Bonferroni-adjusted significance (P < 2.69 × 10^−6^) for at least one combination of variant AF and consequence tier (**Table S6**). In the absence of a well-powered independent replication cohort, we cross-referenced the 11 of 14 candidate CPGs driven by coding variants against a public catalog of gene-based rare coding variant association statistics for 4,529 phenotypes in 394,841 individuals from the UK Biobank (“GeneBass”)^32^, which used comparable variant calling and statistical frameworks. For the 3 candidate CPGs driven exclusively by splicing variants, we also sought supporting evidence from published literature. After review, these approaches prioritized five genes across four cancers that not only surpassed Bonferroni-adjusted significance in our discovery analysis but also exhibited clear convergent evidence in GeneBass or in the literature (**Table 1; Fig. S9**).

**Table 1.** Nominated predisposition genes and associated statistics.

| Gene | Cancer | Criteria | P-value | OR | 95% CI | AF |
| --- | --- | --- | --- | --- | --- | --- |
| *ZNF346* | Kidney | Tiers 1–2 | $4.40 \times 10^{-8}$ | 450.5 | 26.0 – 7,810.9 | 1% |
| *TSTD2* | Thyroid | Tiers 1–4 | $4.86 \times 10^{-7}$ | 111.3 | 11.9 – 1,039.7 | 0.1% |
| *OLFML2B* | Kidney | Tiers 1–3 | $5.46 \times 10^{-7}$ | 78.5 | 9.1 – 674.1 | 1% |
| *CCDC27* | Breast | Tier 1 | $6.29 \times 10^{-7}$ | 1,325.2 | 31.1 – 56,437.9 | 0.1% |
| *BRAT1* | Breast | Tiers 1–2 | $1.05 \times 10^{-6}$ | 10.3 | 3.0 – 35.1 | 1% |
| *MYO1A* | Bladder | Tier 1 | $1.98 \times 10^{-6}$ | 71.8 | 6.1 – 840.1 | 0.1% |

A recurring pattern among these five candidate CPGs was the strong concordance with associations to “proxy” phenotypes reflecting disorders, procedures, or treatments involving the same organ in the UK Biobank cohort. For example, we identified an association between thyroid cancer and rare variants in *TSTD2* in our cohort (Tiers 1–4; P = 4.86 × 10^−7^; OR = 111.3; 95% CI = 11.9–1039.7). In the UK Biobank, *TSTD2* LoF variants exhibited notable associations with “delivery of subsequent element of cycle of chemotherapy for neoplasm” (P = 1.32 × 10^−5^, rank #1 of 4,528 phenotypes tested in GeneBass) and “thyroid surgery” (P = 1.33 × 10^−3^, phenotype rank 13/4,528). In a second example, we identified a strong association between rare variants in *OLFML2B* and kidney cancers (Tiers 1–3; P = 5.46 × 10^−7^; OR = 78.5; 95% CI = 9.1–674.1). GeneBass highlighted a strong association in the UK Biobank between rare LoF variants in *OLFML2B* and “other congenital malformations of kidney” (P = 6.96 × 10^−4^, phenotype rank 4/4,514).

Two of the CPGs nominated in our analyses highlighted contributions of rare noncoding variants predicted to alter splicing, which drove the association signals for these genes. For instance, we identified a rare UTR splice variant (chr7:2555487-C:A) in *BRAT1*, which encodes a BRCA1-associated ATM activator, that was associated with a 10-fold increased risk of breast cancer (Tiers 1–2; P = 1.05 × 10^−6^; OR = 10.3; 95% CI = 3.0–35.1). Although *BRAT1* has not been implicated in breast cancer risk to our knowledge, biallelic LoF variants in this gene are a well-known cause of recessive lethal neonatal rigidity and multifocal seizure syndrome^33^, and its known protein-protein interactions with *BRCA1* and *ATM*^34^ suggest a plausible role in breast cancer susceptibility. In kidney cancer, we identified a rare intronic variant, chr5:177064408-C:G, predicted by SpliceAI^24^ to disrupt splicing of *ZNF346* (Tiers 1–2; P = 4.40 × 10^−8^; OR = 450.5; 95% CI = 26.0–7,811). Although *ZNF346*’s role in kidney diseases is unclear, GeneBass analysis of UK Biobank data showed a strong association between rare nonsynonymous variants in *ZNF346* and “renal failure requiring dialysis” (P = 1.01 × 10^−4^, phenotype rank 1/4,529), supporting its link to kidney disease, potentially including cancer.

Similar to well-established CPGs that predispose to multiple cancer sites, many carriers of rare variants in these candidate CPGs showed incomplete concordance between their cancer diagnoses and their affected FDRs. We hypothesized that subsetting our analyses to cases with concordant family history while adjusting for cancer-specific PRSs would increase power to discover novel CPGs^35^. We therefore defined concordant family history subsets for four cancer types: colorectal and lung cancers (≥1 affected FDR), and breast and prostate cancers (≥2 affected FDRs). This approach identified one additional candidate CPG: rare LoF variants in *CCDC27* (Tier 1; P = 6.29 × 10^−7^) were enriched in breast cancer cases with concordant family histories, with a corresponding GeneBass phenotype, “other disorder of breast” (P = 7.5 × 10^−5^; phenotype rank 1/4,529).

### Runs of homozygosity at specific genomic loci contribute to recessive risk for hereditary cancers

Motivated by known recessive cancer syndromes such as Bloom syndrome and prior work implicating large segments (or “runs”) of homozygosity (RoH) in cancer^36,37^, we next investigated whether RoH might implicate new recessive loci in unexplained familial cancers. We quantified RoH segments spanning ≥100kb and ≥14 common biallelic SNVs (1%< AF < 99%) across all 22 autosomes for each sample. Median genome-wide coverage per sample was 18% for RoHs ≥10 kb, dropping to just 0.3% for very long RoH segments ≥3 Mb, consistent with long RoHs reflecting recent shared ancestry. Finally, we performed genome-wide RoH association scans at four length cutoffs (0.1, 0.25, 0.5, and 1 Mb) across sliding windows of 100 and 250 kilobases, yielding 27,253 approximately independent tests, corresponding to a Bonferroni-adjusted significance threshold of P<1.83×10^−6^. We calculated the proportion of each window covered by RoH per sample and tested associations with each of 18 cancer types using logistic regression adjusted for sex and ancestry (**Table S7**).

Due to substantial variability in haplotype structure across ancestries, our RoH analysis was initially restricted to individuals of European ancestry. Five loci reached Bonferroni-adjusted significance in the RoH discovery analysis, and four remained significant upon inclusion of non-European ancestry groups, with three demonstrating convergent evidence from rare variation in the UK Biobank (**Table 2**; **Fig. S10**). The most compelling association was for RoH ≥100kb at chr22:32,200,001– 32,450,000 with kidney cancer (P = 1.12 × 10^−6^; OR = 9.2; 95% CI = 3.8–22.5), which spans five genes (**Fig. 4a-d**). While most genes in the region lacked associations with renal diseases, rare nonsynonymous variants in *BPIFC* were associated with ‘kidney/renal cell cancer’ (P = 4.88 × 10^−4^, phenotype rank 6/4,529) and ‘nephrectomy’ (P = 1.08 × 10^−3^, phenotype rank 10/4,529) in the UK Biobank^32^. Beyond the chromosome 22 association, we identified two additional genome-wide significant loci. One locus at chr17:30,150,001–30,400,000 was significant in thyroid cancer and contains *NSRP1*, which showed a strong association with “other specified diagnostic endocrinology” in the UK Biobank (P = 8.06 × 10^−5^; phenotype rank 2/4,498). We identified a third RoH association at chr12:22,150,001–22,250,000 in bladder cancer which lies entirely within *ST8SIA1*, a gene associated with “frequency of inability to cease drinking” in the UK Biobank (P = 4.68 × 10^−5^; phenotype rank 1/3,791); this locus may reflect genetic influences on alcohol use, a potential environmental risk factor for bladder cancer^38^, rather than a direct biological contribution to bladder cancer risk. Thus, while the precise inheritance mechanisms remain unclear, these examples demonstrate that RoH at specific genomic loci may recessively contribute to cancer risk in individuals with extensive family history.

**Table 2.**
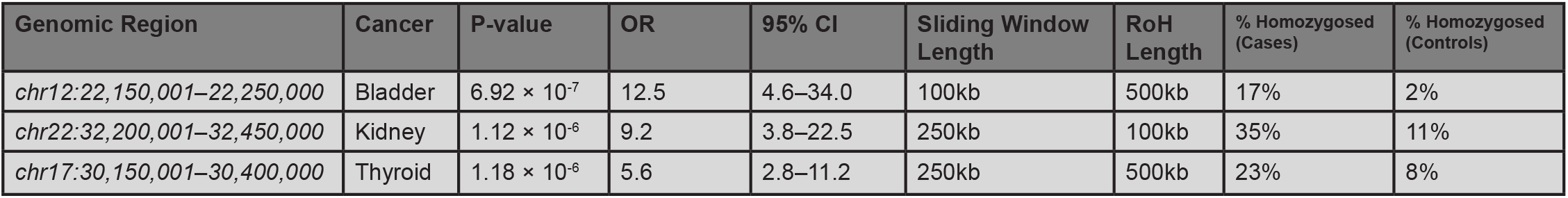
Runs of homozygosity (RoH) loci and associated statistics.

**Figure 4.**
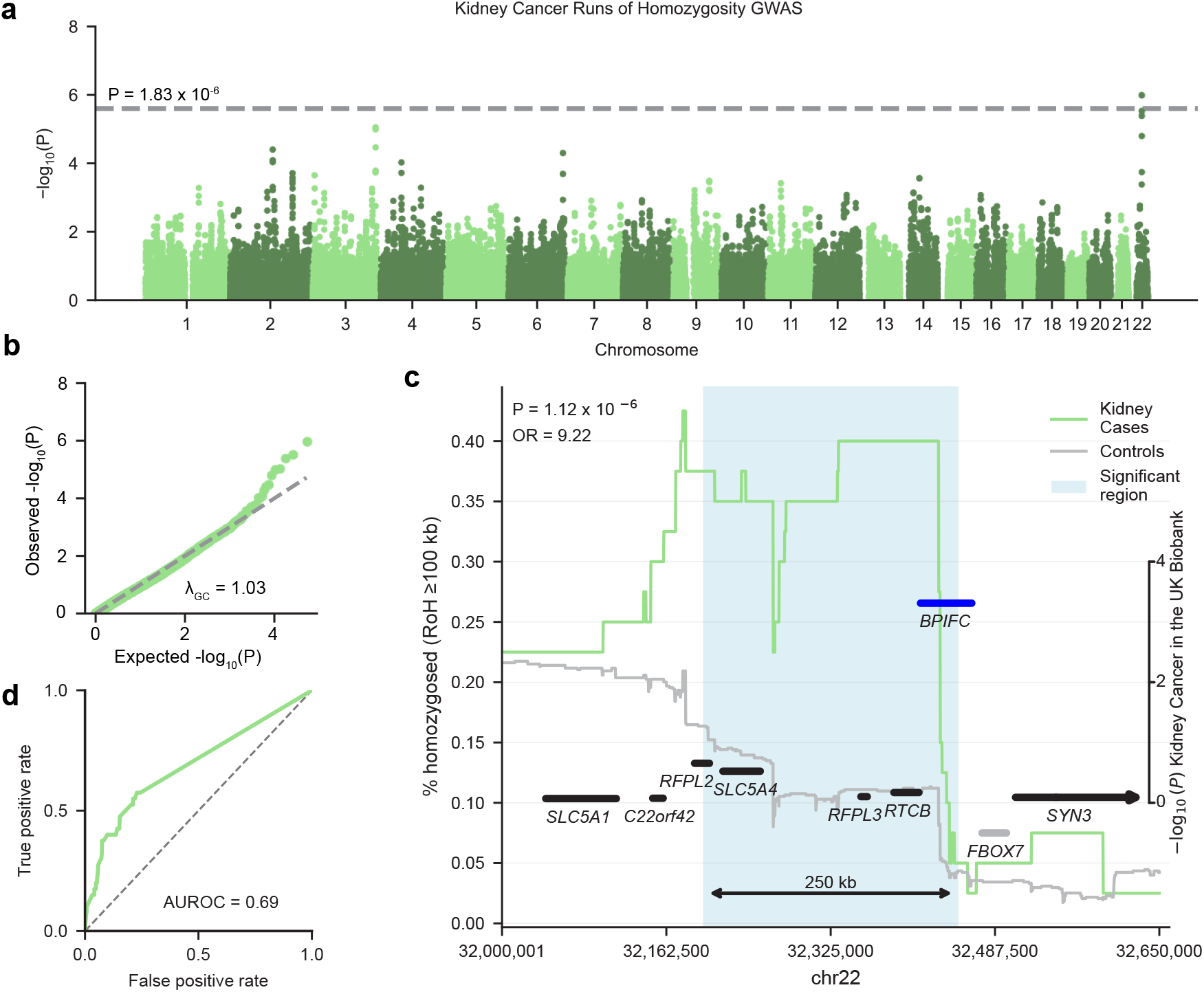
Genome-wide runs of homozygosity (RoH) analyses implicated a candidate recessive kidney cancer predisposition locus involving BPIFC on chr22. **(a)** Results from genome-wide RoH association testing for kidney cancer cases versus controls (RoH ≥ 100kb and 250kb sliding windows). Just one locus on chr22 exceeded Bonferroni significance (P < 1.83 × 10-6). **(b)** Our RoH association models exhibited strong calibration (e.g., kidney cancer shown here). **(c)** Genomic context of the chr22 RoH association with kidney cancer discovered in our analyses. Genes are vertically displayed based on their association with kidney cancer in the UK Biobank for the criteria of missense and low-confidence LoF variants. Genes shown in gray had no recorded phenotypes in the UK Biobank. Individuals with complete homozygosity across the highlighted locus have 9.22-fold higher odds of kidney cancer compared to those with no overlapping RoH ≥100kb. **(d)** RoH coverage (RoH ≥ 100 kb) in chr22:32,200,001–32,450,000 moderately discriminated kidney cancer case–control status in this cohort (AUROC = 0.69), suggesting RoH at loci such as this may be worth evaluating as part of future risk prediction models.

### Estimating the total contribution of germline risk factors to unexplained familial cancers

Having examined myriad genetic predisposition factors across cancer types, we lastly wanted to quantify their cumulative contribution to cancer risk in this unexplained familial cancer population. We therefore computed observed and prevalence-adjusted phenotypic variance explained for each of 18 cancer types using multivariate population attributable fraction regression, incorporating rare LoF SVs and SNVs/indels in cancer type-matched CPGs, PRS, nominated CPGs, and significant RoH loci. We first fit baseline logistic regression models per cancer, including just two demographic covariates, sex and ancestry, which explained 0.6–7.1% (median = 3.2%) of risk across 18 cancer types. Next, we extended these regression models by incorporating risk factors involving known cancer-associated loci, namely PRS and rare variants in cancer type-matched CPGs. We found that these genomic risk factors increased phenotypic variance explained by a median of +4.6% (range: +0.4–18.1%) in our cohort (**Fig. 5a**), which translated to an increase of +0.5–9.8% of liability-scale heritability after adjusting for estimated population prevalence of unexplained familial cancers^39–41^. Across most cancer types, PRS accounted for the largest gain among any single category of risk factors (mean = +4.7% due to PRS alone), while the proportion of variance explained due to rare nonsynonymous variants in CPGs often correlated with concordant family history (**Fig. 5b-c**). Finally, we extended these regression models by incorporating all new candidate risk factors discovered in this study, including (i) 6 new CPGs and (ii) 3 significant RoH loci. After accounting for the variance explained by the baseline models and established loci, the new risk factors we nominated in this study explained an additional +2.4–18.5% (median = +6.2%) of cancer risk among unexplained familial cases for the four cancer types in which we identified at least one new candidate risk factor (**Fig. S11**). The variance explained by new risk factors corresponded to an average increase of +10.0% when adjusting for population prevalence of familial cancers. In total, these population attributable fraction calculations estimated that the totality of all risk factors considered in this study likely provided an explanation for between 1.0% (brain) and 25.0% (kidney) of all unexplained cases of familial cancers across the 18 cancer types evaluated here, firmly demonstrating the value of germline whole-genome sequencing in genetic studies of families with unusually high clustering of cancers.

**Figure 5.**
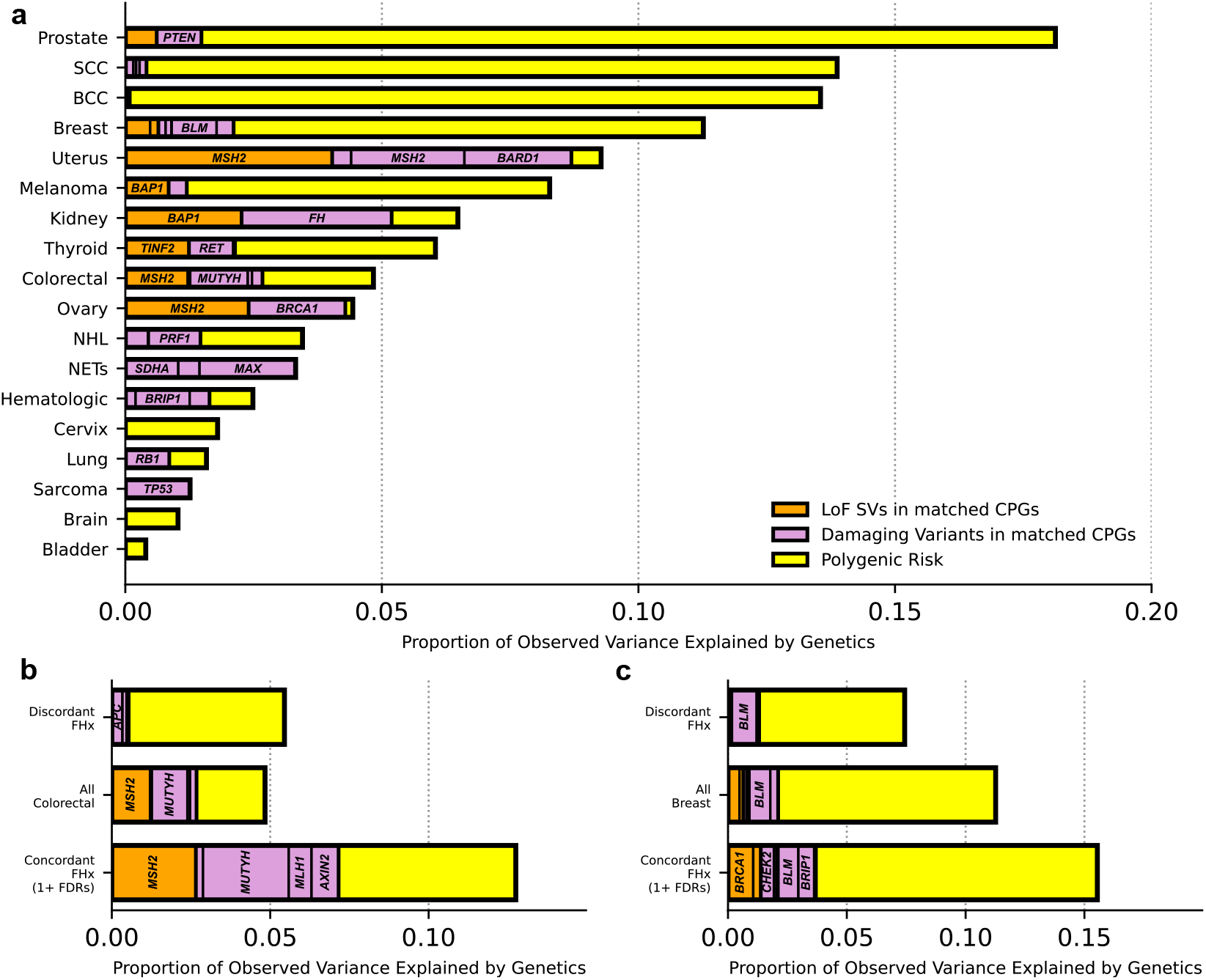
The contribution of germline genetic factors beyond canonical PGVs to familial cancer liability. **(a)** Proportion of observed phenotypic variance explained across 18 cancer types in our study when incorporating underappreciated classes of variation involving known cancer risk loci: LoF SVs in five selected genes; rare SNVs/indels in cancer type-matched CPGs; and PRS. Proportion of observed variance explained in **(b)** colorectal cancer and **(c)** breast cancer when stratifying by concordant family history.

## DISCUSSION

Family history is a major risk factor for cancer, yet only a minority of cancer patients ascertained with positive family history exhibit a clinically recognized genetic predisposition. In this study, we aimed to close this gap by comprehensively evaluating the genomic architecture of unexplained familial cancers. We systematically analyzed rare and common variation— including coding and noncoding variants, SNVs, indels, and SVs—across 2,726 germline whole genomes from a large national biobank to better understand the inherited basis of unexplained familial cancers.

The overarching conclusion from our study is that familial cancer risk reflects a continuum of genetic mechanisms beyond canonical PGVs, spanning rare LoF SVs, elevated polygenic risk, and homozygosity of common variation, nearly all of which appear to act as incompletely penetrant risk modifiers rather than Mendelian dominant loci. This observation diverges from traditional models of familial disease wherein phenotypic segregation within a pedigree is often assumed to track with a single dominant or recessive risk factor, as is the case for most well-known cancer predisposition syndromes, like Lynch syndrome. In particular, our PRS analyses demonstrated that high cancer PRS concentrated within a family can cause apparent quasi-Mendelian familial cancer clustering, and that the unequal inheritance of high PRS between family members likely contributes to the distribution of specific cancers within families.

Our biobank WGS-based strategy was also successful in nominating six new candidate CPGs, including two novel CPGs in breast cancer: *BRAT1* and *CCDC27*. Given the exhaustive prior gene discovery efforts in breast cancer^42,43^, it may seem surprising that our relatively modest cohort was able to nominate two new genes; however, both *BRAT1* and *CCDC27* highlight situations that were uniquely accessible to our combined WGS analyses and cohort ascertainment due to being driven by a single rare UTR splice variant inaccessible to technologies like microarrays or exome sequencing (*BRAT1*), or only emerging as significant after enforcing concordant family history and adjusting for each individual’s breast cancer PRS (*CCDC27*). We further identified allelic series of LoF SVs affecting *MSH2* and *BRCA1* in patients with phenotypes consistent with Lynch syndrome and hereditary breast and ovarian cancer syndrome, underscoring that rare SVs impacting known CPGs may contribute to unexplained hereditary cancers. Finally, we also discovered three loci where RoH of common variants were significantly associated with cancer risk, all of which colocalized with a plausible candidate causal gene (*BPIFC, ST8SIA1*, and *NSRP1*), although further work is needed to determine the precise mechanism at each locus.

The design of our current study cohort allowed us to heavily enrich cases for novel genetic mechanisms of cancer risk, but this bespoke ascertainment criteria also resulted in significant limitations, including challenges in replicating these findings in comparable cohorts. Additionally, our strict ascertainment criteria produced modest sample sizes even without strictly matching cancer types between affected probands and their FDRs, which limited our statistical power compared to larger, unselected case–control designs. By requiring controls to report no family history of cancer and thus depleting their genetic cancer risk, we anticipate that most effect size estimates reported here are inflated compared to their true effects in the general cancer patient and presymptomatic at-risk populations, potentially limiting the direct generalizability of our discoveries. For example, our final population attributable fraction estimates are likely upper bounds of the true contribution of novel risk factors due to Winner’s Curse and therefore should be taken as coarse projections rather than precise expected diagnostic yields. We note other limitations beyond cohort ascertainment: for example, our analyses of known CPGs did not consider differences in inheritance patterns (recessive vs. dominant) or gene function (tumor suppressor vs. oncogene), even though many CPGs act recessively or through molecular mechanisms other than loss-of-function. Lastly, reliance on All of Us EHR and survey data limited phenotypic resolution, including detailed histologic subtypes, comorbidities, environmental covariates, and access to matched tumor sequencing. Larger WGS studies of cancer patients with paired tumor profiling and deeper clinical annotation will be essential to confirm and extend these findings.

Overall, this study provided one of the most comprehensive pan-cancer evaluations of genomic risk mechanisms in familial cancers beyond PGVs to date. Our results indicate that several cancers, such as kidney and thyroid cancer, may have a larger inherited component than previously appreciated. Ultimately, the progressive incorporation of higher resolution genomic methods like WGS into clinical settings is likely to improve risk stratification, personalized screening, and new prevention strategies for cancer-burdened families.

## Data Availability

All data produced in the present study are available upon reasonable request to the authors

## ACKNOWLEDGMENTS

We gratefully acknowledge the *All of Us* Research Program’s participants for their contributions, without whom this research would not have been possible. We also thank the National Institutes of Health’s *All of Us* Research Program for making available the participant data and genome sequencing examined in this study. We also would like to thank the members of the Van Allen Lab for their thoughtful comments and suggestions throughout the course of this study.

## Author Contributions

Conceptualization: N.F., S.H., J.P., R.G., R.L.C., E.M.V.A; Formal analysis: N.F., R.L.C; Funding acquisition: E.V.M.A., R.L.C.; Investigation: N.F., R.L.C; Methodology: N.F., R.L.C.; Resources: E.V.M.A., R.G., S.H.A., J.K., J.G., D.N., R.B., E.S.; Writing - original draft: N.F., R.L.C; Writing - review and editing: E.M.V.A., E.P., J.P., R.G., S.H.A, S.H, J.G., W.M.

## METHODS

### Sample Selection and Cohort Creation

The genomic and phenotypic data presented in this study were sourced from the All of Us Research Program Curated Data Repository version 7 (CDR v7)^13,14^. We restricted the totality of participants from All of Us CDR v7 to those with available short-read germline WGS, electronic health records (EHR), and self-reported survey data for family history of cancer. We defined familial cancer cases as participants with at least three cancer-affected first- or second-degree relatives (defined as grandparent, mother, father, sibling, son, daughter, or grandchild). Notably, the All of Us dataset only records the presence of a relative type with cancer and does not capture multiple affected individuals within the same category (e.g., it does not distinguish between one vs. multiple siblings with cancer). This strict filtering yielded 2,860 potential cases. We next leveraged existing germline exome SNV/ indel calls^13^ for these individuals, through the All of Us Controlled Tier, to exclude those harboring potential germline predisposition variants in any of 148 cancer predisposition genes (CPGs). Specifically, we excluded individuals with ClinVar Pathogenic/Likely Pathogenic variants or rare (AF < 1%) *HIGH*-impact variants, defined by the Variant Effect Predictor^44^, in a CPG, resulting in a final set of 1,496 cases. For simplicity, we collectively dubbed these variants pathogenic germline variants (PGVs). Notably, rare *HIGH*-impact variants that were annotated in ClinVar as Benign/Likely Benign were not included in this set of PGVs. We derived our list of 148 CPGs from a recent rare variant study of pediatric sarcomas (**Table S1**). We intentionally erred towards conservatively excluding samples when applying this PGV SNV/indel filter by considering all transcripts of CPGs defined by Gencode v44^45^ and applying no additional filters to *HIGH*-impact variants, so we likely overestimated the number of individuals containing truly pathogenic LoF variants in CPGs. Reciprocally, we defined cancer-free controls as individuals who had no cancer diagnosis and no relatives reported with cancer. We further matched controls to cases by age, sex, and reported ethnicity. This initially left us with 3,224 individuals, however after variant calling, joint genotyping, and variant quality control, we decided to also (i) use the most recent version of ClinVar pathogenicity annotations to further exclude individuals who no longer met our filtering criteria, and (ii) ensure controls also carried no PGV SNV/indel in CPGs as defined above. In total, these phenotypic and genomic filters yielded 2,726 individuals, including 1,496 cases and 1,230 controls.

### Cancer phenotype curation

We next assigned categorical cancer diagnosis labels for all 1,496 cases prioritized by our filtering above. Due to limitations inherent in EHR data, cases were grouped by histology only where possible without introducing significant ambiguity; in most instances, grouping was based on the cancer’s organ of origin rather than specific histological type. For example, diagnoses of “primary malignant neoplasms of the kidney” were labeled as “kidney cancer” regardless of histological subtype. We provide a full mapping of EHR terms to cancer diagnosis labels used in this study in **Table S2**. These mappings were cross-evaluated by multiple board-certified medical oncologists to ensure approximate consistency with current clinical knowledge. We applied additional filters to the provided diagnostic codes to reduce heterogeneity while retaining sample size where possible:

1. Lung cancer cases did not include tumors of pleural origin and lung carcinoid tumors, though respiratory tract tumors were included;
2. Cervical cancer cases included both invasive and intraepithelial neoplasms;
3. Breast cancer cases were restricted to females. There were ≤20 males diagnosed with breast cancer in our cohort and were therefore not included per All of Us’s data and statistics dissemination policy;
4. Bladder cancer cases included cancers of the renal pelvis and ureter due to their shared urothelial origin, whereas kidney cancer cases excluded renal pelvis tumors;
5. Hematologic cancer cases included leukemias, mast cell neoplasms, and myelomas;
6. Melanoma cases excluded ocular melanomas;
7. Medullary thyroid carcinomas were classified as neuroendocrine tumors rather than thyroid cancers.

When generating the 18 cancer cohorts used in this study, we first excluded all individuals with inferred sex chromosome ploidy other than XX or XY (n ≤ 20). We then pruned all pairs of individuals with a kinship coefficient ≥0.1 by retaining the individual with the earliest onset of any cancer type from each pair to exclude cryptic relatedness (n ≤ 20 individuals with a related individual in this study). We then matched controls to our cases using sex and continental ancestry generated from GRAF-pop^46^ and subsequent clustering to the 1000 Genomes Project^47^ reference populations. Specifically, we downsampled a maximally large set of controls for each of 18 cancer types while matching controls to cases on proportions of continental ancestry and genetic sex. Control selection was further refined by matching the age distributions of controls to those of the cases. In accordance with the All of Us data and statistics dissemination policy, counts or percentages for fewer than 20 individuals are not reported.

### Matching cancer predisposition genes to COSMIC cancer types

We manually matched our list of 148 CPGs to cancer types annotated as exhibiting a germline association per the COSMIC Cancer Gene Census^25^. This filtering retained 79 genes that were listed as CPGs for at least one of the 18 cancers analyzed in this study. This refined map of 79 genes to our 18 cancer types was used only (i) to analyze the enrichment of rare SNVs/indels in CPGs (**Fig. 2f-h**) and (ii) to calculate population attributable fraction due to rare SNVs/indels in CPGs (**Fig. 5a-c; Fig. S11**). Because COSMIC cancer types often reflect highly specific histologies rather than the coarse organ-level diagnoses we curated from the All of Us EHR, we mapped a minority of detailed COSMIC entries to broader cancer types; for example, we matched COSMIC’s asserted association of germline *STAT3* variants in “pediatric large granular lymphocytic leukemia” to the “hematologic cancer” cancer type in our All of Us cohort. We manually curated cancer associations for *BLM, TP53*, and *WRN*^48–50^, as the germline annotations in COSMIC lacked specific, well-known cancer associations (e.g., rare LoF variants in *WRN* as a predisposition factor to thyroid cancer and melanoma).

### Germline short variant discovery and genotyping

We performed germline SNV and indel discovery with the Genome Analysis Toolkit (GATK) jointly from germline WGS for all qualifying individuals from All of Us CDR v7, plus select multi-sample pedigrees from the Centre d’Etude du Polymorphisme Humain (CEPH)^51,52^, Dana Farber Cancer Institute (DFCI)^53^, and the Multi-Ethnic Study on Atherosclerosis (MESA)^54,55^. The inclusion of these families from non-All of Us cohorts enabled rigorous variant quality control and filtering (e.g., by analyzing rates of de novo variants) that would not have been possible without including these benchmarking datasets. We thus started with 4,769 WGS samples (599 CEPH, 899 MESA, 47 DFCI, 3,224 All of Us)^14,52,54^.

We began by applying GATK HaplotypeCaller (GATK-HC) version v4.2.0.0 to all 4,769 WGS samples to produce single-sample gVCFs^56^. We next performed an initial permissive quality control of short-read WGS data, excluding 131 samples based on unusually high (>45x) or low (<15x) coverage, sample contamination (Charr > 0.03)^57^, exceptionally biased WGS coverage^17^, or poor GATK-HC calling metrics leaving us 4,638 WGS samples (539 CEPH, 834 MESA, 47 DFCI, 3,218 All of Us). As described in the “Sample Selection and Cohort Creation” section, these 3,218 All of Us samples were further (after variant quality control) reduced to 2,726 by excluding individuals based on updated ClinVar pathogenicity annotations and by applying the same exclusion criteria to controls, which were not initially filtered for the presence of PGVs in the 148 CPGs. We next jointly genotyped all SNV/indel sites across all samples using the new GATK “Biggest Practices / Gnarly” pipeline developed for the Genome Aggregation Database (gnomAD) v3.0^58^. Following joint genotyping, we hard-filtered all raw variant calls based on minimum read depth (DP) ≥10, genotype quality (GQ) ≥20, and variant allele fraction (VAF) ≥0.2.

### Short variant filtering and quality control

To further reduce false-positive variant calls, we trained a Random Forest classifier on the raw variant set, following the methodology established in gnomAD^59^. As these methods are previously described and publicly accessible (http://gnomad-sg.org/help/variant-qc, https://broadinstitute.github.io/gnomad_methods/_modules/gnomad/variant_qc/random_forest.html), we provide only a brief overview of the input to the Random Forest approach here. We defined likely true positive variants (TPs) using high-confidence variant sets from public resources and internal data. Specifically, we included SNVs present in the Omni 2.5 genotyping array and in the 1000 Genomes Project^60^, as well as indels from the Mills and Devine dataset^61^. In addition, we incorporated transmitted singletons, defined as variants observed in exactly two individuals corresponding to a parent-offspring pair within the CEPH, MESA, and DFCI cohorts. Likely false positive variants (FPs) were defined as those failing traditional GATK hard filters (AS_QD < 2, AS_FS > 60, AS_MQ < 30). To train our Random Forest model, we balanced the number of TPs and FPs to create a 50:50 split of training variants. All variants were subsequently annotated using multiple tools: GATK v4.6.0.0 was used to assign Variant Type and classify indels as h-indel or non-h-indel, and GATK v3.8 was used to calculate AS_ Inbreeding_Coeff, leveraging pedigree data from the CEPH, MESA, and DFCI cohorts. We employed Hail v0.2.127^62^ to annotate each variant with pab_max_expr. The Variant Type category consisted of SNV, non-h-indel, and h-indel. We trained a random forest classifier by using the code (URL cited above) used for gnomAD v2.1. We made slight adjustments to the methods as suggested by their website and calculated certain metrics using allele-specific metrics as opposed to site-specific metrics (i.e., Inbreeding Coeff, MQRankSum, ReadPosRankSum, SOR).

We applied the trained random forest classifier to assign a true positive probability (TPP) to each variant. Thresholds were selected to (i) minimize de novo variants in the CEPH, MESA, and internal DFCI trios, (ii) ensure total variant count aligned with expected count based on prior studies, (iii) to limit variants that did not align with Hardy–Weinberg equilibrium (HWE), and (iv) to maximize sensitivity and overall variant retention relative to high-confidence SNVs from the 1000 Genomes Project (phase 1) and indels from the 1000 Genomes Project and Mills gold standard datasets (version 0). When analyzing HWE statistics per variant, we analyzed only common (AF > 1% and > 5%) biallelic variants in individuals who most closely matched European ancestry groups in the All of Us cohort. HWE p-values per variant were calculated using VcfTools v0.1.15^63^. When performing *de novo* variant counts for probands in trios (available only for CEPH, MESA, DFCI), we excluded variants with ALT alleles marked as “*” or in low-quality regions enriched for segmental duplications. We also excluded variants with cohort AF ≥0.1%, read depth ≤10, or genotype quality ≤20. The final TPP thresholds (0.63 for SNVs and 0.76 for indels) were selected based on an optimal balance of these metrics, including variant sensitivity, HWE, de novo variant minimization, and total variant count (**Fig. S2, S3**).

### Genetic ancestry and relatedness inference

We determined genetic ancestry for all samples by analyzing common SNVs that passed the post hoc random forest filtering described above. We first filtered all SNVs to retain variants also present in the 1000 Genomes Project^47^ and had AFs between 1% and 99% with at least 100 non-reference alleles called, and a quality-by-depth score >10. We further restricted to biallelic SNVs in Hardy–Weinberg equilibrium (P > 0.05). We next pruned SNVs to retain unlinked variants by performing linkage disequilibrium (LD) pruning with PLINK2^64^ using 10 Mb sliding windows, removing one SNV from each pair with r^2^ >0.1 and advancing the window in steps of 5 SNVs. Principal component analysis (PCA) was conducted with PLINK2 to capture major axes of ancestry-correlated genetic variation using only remaining SNVs with genotype call rates >95%. Pairwise kinship coefficients were also estimated from these same filtered SNVs using PLINK2 with the --make-king-table flag, and sample pairs with kinship >0.1 were flagged using --king-table-filter 0.1.

### Joint SV discovery, genotyping, and filtering

We produced a genome-wide SV callset for all WGS samples using GATK-SV, a cloud-based pipeline tailored to large-scale WGS cohorts^17^. Each WGS sample was first processed according to the recommended default configuration of GATK-SV v1.0.1. Next, as short-read WGS-based SV calling performance improves with larger cohort sizes, all samples described in this study were joint-called with GATK-SV alongside 53,959 other germline WGS samples that were part of other unrelated & ongoing studies and institutional initiatives. We applied GATK-SV in cohort mode to integrate and jointly genotype all putative SV calls across all WGS samples into a single, unified SV callset.

After executing the GATK-SV pipeline, we next applied numerous established and custom filters to boost the specificity of this SV callset. We began by excluding (i) unresolved SVs, or “breakends”, (ii) deletion SVs detected exclusively by the Wham algorithm, and (iii) SVs that did not have at least one non-reference genotype carrier with genotype quality (GQ) > 1. Following these hard filters, we applied a series of methods similar to our prior WGS study of pediatric cancers^65^, which began by applying a pre-trained SV GQ recalibration model that was produced by the All of Us SV production team^13^. Given that our current cohort was also derived from a subset of the same All of Us WGS data, we reapplied this pre-trained GQ recalibration model exactly as provided with default parameters. We filtered SV genotypes using the All of Us-recommended “high-sensitivity” GQ thresholds and excluded all SVs with a call rate <90% after masking low-quality genotypes. Next, we applied a trio-based filtering method that we previously developed for gnomAD, which trained a decision tree of GQ filters optimized towards a 5% target Mendelian violation rate in parent-child trios from the ancillary CEPH, MESA, and DFCI samples included for quality control (see above) following our previously published protocol^17^. We then applied our established methods to exclude predicted genic retrocopy insertions, common insertions incorrectly resolved by GATK-SV as reciprocal translocations, and mask predicted batch-or cohort-specific effects^65^. Following all SV filtering, we identified and excluded SV-specific outlier samples defined as genomes that carried a total number of SVs from any one mutational class that exceeded the third quartile plus three times the interquartile range across all samples for that same SV class; this analysis was conducted while restricting within each continental ancestry group to account for known ancestry-correlated differences in SV burden.

### Functional consequence annotation

We annotated the functional consequences of SNVs/indels with the ENSEMBL Variant Effect Predictor (VEP)^44^ v115 for variants ≤50bp. Missense variants were further annotated using REVEL^22^ and PrimateAI^23^ via dbNSFP version 3a for GRCh38^66^, as well as with AlphaMissense^21^. We classified missense variants as predicted damaging or weakly damaging (**Fig. 2f**) based on the number of in silico models predicting a deleterious effect (damaging: ≥2 of 3; weakly damaging: 1 of 3). Predicted splicing effects were classified as splice-disruptive if SpliceAI v1.3^24^ reported a score of ≥0.5 for any category (i.e., acceptor gain, acceptor loss, donor gain, donor loss; **Fig. 2f**). VEP-predicted LoF variants were further annotated by the Loss-of-Function Transcript Effect Estimator (LOFTEE)^59^. Additionally, we downloaded ClinVar’s^20^ October 19, 2025 VCF update and used its variant classifications (e.g., “Pathogenic,” “Variant of Uncertain Significance”) for our pathogenicity (tier) classifications (**Fig. S8**). In parallel, we annotated all SVs with GATK SVAnnotate to assign no more than one of 11 predicted functional consequences for each gene impacted by each SV^17^. We used the Gencode v47 basic gene annotation file for all genic SV predictions^45^. For the purposes of this study, we defined an SV as LoF if it was assigned the LoF, intragenic exonic duplication, or partial exon duplication consequence by GATK.

### Variant frequency definitions

Throughout this study, we applied various AF thresholds to isolate rare variants. The exact definitions of these thresholds are defined here. We define rare SNVs/indels as those with AF <1% in African, Non-Finnish European, East Asian, South Asian, American, and Finnish non-cancer populations in gnomAD v3.1^59^, as well as in our cohort from All of Us. To account for bottlenecked or undersampled populations in gnomAD, we relaxed these criteria to AF <10% for Middle Eastern, Amish, Ashkenazi Jewish, and “Other” populations in gnomAD v3.1. We defined SNVs/indels as “ultra-rare” if they exhibited AF <0.1% in the same larger non-cancer populations (i.e., African, Non-Finnish European, East Asian, South Asian, American, Finnish, and in our All of Us cohort) and <1% for the smaller populations in gnomAD (i.e., Middle Eastern, Amish, Ashkenazi Jewish, and Other). Rare SVs were defined using WGS-based SV frequencies from gnomAD v4.1^59^, applying the same population-specific AF thresholds as for SNVs/indels. Rare variants for quality control and count data were obtained using an AF of 1% within our 2,726 individuals analyzed here.

### Computing ancestry-controlled polygenic risk scores

We computed 39 polygenic scores across four published studies that supplied genome-wide weights for multiple cancer types^26–29^. While most PRS models were trained on cancer types directly corresponding to those analyzed in our study, in some cases, we used closely related phenotypes as proxies (e.g., lymphocytic leukemia PRS for hematologic cancers and glioblastoma PRS for brain cancers). We assessed a total of 39 polygenic risk models by applying the PLINK v1.9 score function^64^ to project each sample’s genotypes through the published PRS weights. To adjust for potential confounding effects seen in multi-ancestry PRS studies^67^, these raw PRS values were regressed for sex and the top 4 genetic principal components using linear regression, yielding residual PRS values. We subsequently standard-normalized these residual PRS values against cancer-free, covariate-matched controls as a baseline to a mean of 0 and a standard deviation of 1. These resulting adjusted PRS values were subsequently used to evaluate the discriminatory power of each score between cases and controls. This entire procedure was repeated while restricting to individuals of European ancestry in each of the 14 cancer types evaluated in our study with a corresponding PRS model. Although we evaluated each of the 39 PRS models on their respective cancer types (e.g., breast cancer PRS in breast cancer cases vs. controls), subsequent analyses were restricted to the most significant PRS model per cancer type, defined as the model with the lowest p-value in its corresponding cancer type. This procedure retained 14 PRS models for further analysis after excluding the four cancer types for which no matching PRS was available.

To minimize confounding when evaluating a PRS across mismatched cancer types (*e*.*g*., breast cancer PRS model applied to lung cancer cases), we excluded cases diagnosed with both the PRS-matched cancer and the cancer under analysis. Similarly, when evaluating the relationship between family history and PRS, we generated a new set of controls per cancer type to match all cases with a family history of a specific cancer, ignoring that patient’s actual diagnoses (**Fig. 3d**; **Fig. S7**).

### Gene-based rare variant association testing

We performed exome-wide rare variant collapsing burden association tests using SAIGE-GENE+ following its recommended procedures.^31^ We first created a sparse genetic relatedness matrix using a linkage-disequilibrium pruned, common, biallelic SNVs filtered from the 1000 Genomes Project^18^. We then used the sparse genetic relatedness matrix to train the SAIGE-GENE+ null model. Covariates included genetic principal components 1–4 and sex. When performing the gene discovery analysis in individuals with a concordant family history, we incorporated the most significant cancer type-matched PRS as a covariate. We tested 18,544 autosomal protein-coding genes annotated in Gencode v47^45^ across all 15 cancer types with ≥40 cases in our cohort. For each gene, we further considered four variant impact criteria and two AF thresholds (1% and 0.1%). Loss-of-function (LoF) associations were analyzed incrementally, starting with Tier 1 variants and sequentially adding Tiers 2–4 (**Fig. S8**). A tradeoff of the SAIGE-GENE+ method when executed with default parameters is that it performs AF-based weighting of variants per gene to boost discovery power; however, due to this variant weighting, the default implementation of SAIGE-GENE+ cannot produce realistic estimates of effect sizes for genes. Thus, to improve effect size estimation, we executed SAIGE-GENE+ twice: first to produce maximally powered test statistics to define cancer-associated genes, and a second time while disabling variant reweighting to estimate effect sizes for all genes. Finally, p-values for each pathogenicity–AF combination were recalibrated using saddlepoint approximation to account for case– control imbalances using a similar implementation as we have previously published^65,68^.

### Runs of homozygosity analysis

We performed genome-wide detection of runs of homozygosity (RoH) in all genomes. To accomplish this, we first restricted the dataset to common, biallelic SNVs with AF between 1% and 99%, a minimum quality depth (QD) >10, Hardy–Weinberg p >0.01, and at least 100 alleles detected in the cohort. After variant filtering, we ran *bcftools roh*^69^ on all autosomes using a reference genetic map^70^ to call RoH. Because *bcftools roh* can redundantly call certain loci, each sample’s RoH was merged with bedtools^71^ to ensure no genomic loci were double counted.

We subsequently performed genome-wide RoH association scans at four minimum RoH length cutoffs (0.1, 0.25, 0.5, and 1 Mb) across sliding windows of 100 and 250 kilobases, yielding 27,253 approximately independent tests (calculated by using non-overlapping 100kb windows in the 22 autosomes, excluding windows within 2Mb of centromeres^72^), corresponding to a Bonferroni-adjusted significance threshold of P<1.83×10^−6^. Sliding windows of all lengths were generated using 50 kb increments. We calculated the proportion of each window covered by RoH per sample and tested associations with each of 18 cancer types using logistic regression adjusted for sex and ancestry. Genome-wide association analyses were restricted to windows where at least one case and one control had a RoH overlapping the region, evaluated separately for each cancer type. We collapsed overlapping significant loci identified at different test resolutions and reported the window spanning more base pairs, breaking ties by selecting the most significant p-value among all tests at that locus.

### Population attributable fraction analysis

We estimated the incremental contribution of sequentially added predictors (genetic risk factors) to risk for each of 18 cancer types using logistic regression-based attributable fraction analyses. Each model included a fixed set of baseline covariates consisting of the first four genetic principal components, sex, and a binary indicator for whether each sample passed SV-specific quality control. Predictors of interest were evaluated in a pre-specified order: LoF SVs of 5 selected genes (described below), rare SNVs/ indels in cancer-matched CPGs, PRS, nominated CPGs, and nominated RoH loci. Models were constructed sequentially by cumulatively adding predictors one at a time. For each added predictor, we fit a full logistic regression model including all covariates and the current set of predictors, as well as a reduced model including only covariates.

While PRS and the presence of potentially pathogenic variants in nominated CPGs (e.g., *BRAT1, TSTD2*) were straightforward to incorporate, the inclusion of LoF SVs, rare variants in cancer-matched CPGs, and nominated RoH loci required additional curation. We restricted LoF SVs to rare events in a subset of well-established CPGs not seen in controls (i.e., *ATM, BAP1, BRCA1, MSH2*, and *TINF2*), and only for cancer types with known gene–disease associations (e.g., *MSH2* for colorectal, uterine, and ovarian cancers, but not squamous cell carcinoma; **Fig. 2a**). For rare SNVs/indels in cancer-matched CPGs, we identified the most significant variant category (defined by functional consequence and AF; **Fig. 2f**) with an OR >1, decomposed the corresponding gene set into individual genes, and modeled the presence of a qualifying rare variant in each gene as a separate predictor. Finally, for nominated RoH loci, we retained only those that remained significant after Bonferroni correction and showed convergent evidence from rare variation analyses in the UK Biobank. We then modeled homozygosity at each locus as a continuous variable ranging from 0 to When multiple Bonferroni-adjusted significant loci overlapped or were adjacent, we prioritized larger loci, with secondary prioritization based on more significant p-values.

Model fit was quantified using Nagelkerke’s pseudo-R^2^ relative to an intercept-only null model. Rather than estimating predictor-specific attributable fractions, we focused on the cumulative contribution of genetic predictors following sequential model adjustment. Nagelkerke’s pseudo-R^2^ estimates were transformed to the liability scale using the method of Lee et al.41, incorporating cancer prevalence estimates from the National Cancer Institute’s Surveillance, Epidemiology, and End Results (SEER) Program^39^. Because SEER prevalence estimates do not account for a family history of cancer, we additionally derived an adjusted prevalence that we reasoned would more accurately reflect familial cancer burden by scaling SEER estimates by the proportion of cases with a concordant cancer diagnosis in a parent or sibling in the Swedish Family-Cancer Database (SFCD)^40^. Using these prevalence definitions, we derived three sets of liability-scale estimates for the cancer types in our study. First, we report observed variance explained by genetic risk factors using Nagelkerke’s pseudo-R^2^. Second, we transformed these estimates to the liability scale using SEER population prevalence. Third, we further refined these estimates by incorporating an adjusted prevalence that accounts for familial cancer enrichment, derived from the SFCD.

## REFERENCES

1. Sundquist, K., Sundquist, J. & Ji, J. Contribution of shared environmental factors to familial aggregation of common cancers: an adoption study in Sweden. Eur. J. Cancer Prev. 24, 162 (2015).

2. Mucci, L. A. et al. Familial Risk and Heritability of Cancer Among Twins in Nordic Countries. JAMA 315, 68–76 (2016).

3. Huang, K. et al. Pathogenic Germline Variants in 10,389 Adult Cancers. Cell 173, 355–370.e14 (2018).

4. Shiovitz, S. & Korde, L. A. Genetics of breast cancer: a topic in evolution. Ann. Oncol. 26, 1291–1299 (2015).

5. Schubert, S. A., Morreau, H., de Miranda, N. F. C. C. & van Wezel, T. The missing heritability of familial colorectal cancer. Mutagenesis 35, 221–231 (2020).

6. Alba-Pavón, P., Alaña, L., Astigarraga, I. & Villate, O. Splicing-Disrupting Mutations in Inherited Predisposition to Solid Pediatric Cancer. Cancers 14, 5967 (2022).

7. Pócza, T. et al. Germline Structural Variations in Cancer Predisposition Genes. Front. Genet. 12, 634217 (2021).

8. Wang, L. et al. Performance of polygenic risk scores for cancer prediction in a racially diverse academic biobank. Genet. Med. 24, 601–609 (2022).

9. Cheng, W. et al. Genome-wide analysis of runs of homozygosity identifies new susceptibility regions of lung cancer in Han Chinese. J. Biomed. Res. 27, 208 (2013).

10. Lappalainen, T., Scott, A. J., Brandt, M. & Hall, I. M. Genomic Analysis in the Age of Human Genome Sequencing. Cell 177, 70–84 (2019).

11. Collins, R. L. & Talkowski, M. E. Diversity and consequences of structural variation in the human genome. Nat. Rev. Genet. 26, 443–462 (2025).

12. Lionel, A. C. et al. Improved diagnostic yield compared with targeted gene sequencing panels suggests a role for whole-genome sequencing as a first-tier genetic test. Genet. Med. 20, 435–443 (2018).

13. The All of Us Research Program Genomics Investigators et al. Genomic data in the All of Us Research Program. Nature 627, 340–346 (2024).

14. Ramirez, A. H. et al. The All of Us Research Program: Data quality, utility, and diversity. Patterns 3, 100570 (2022).

15. Gillani, R. et al. Germline predisposition to pediatric Ewing sarcoma is characterized by inherited pathogenic variants in DNA damage repair genes. Am. J. Hum. Genet. 109, 1026–1037 (2022).

16. McKenna, A. et al. The Genome Analysis Toolkit: a MapReduce framework for analyzing next-generation DNA sequencing data. Genome Res. 20, 1297–1303 (2010).

17. Collins, R. L. et al. A structural variation reference for medical and population genetics. Nature 581, 444–451 (2020).

18. Byrska-Bishop, M. et al. High-coverage whole-genome sequencing of the expanded 1000 Genomes Project cohort including 602 trios. Cell 185, 3426–3440.e19 (2022).

19. Martínez-Roca, A. et al. Lynch-like Syndrome: Potential Mechanisms and Management. Cancers 14, 1115 (2022).

20. Landrum, M. J. et al. ClinVar: improving access to variant interpretations and supporting evidence. Nucleic Acids Res. 46, D1062–D1067 (2018).

21. Cheng, J. et al. Accurate proteome-wide missense variant effect prediction with AlphaMissense. Science 381, eadg7492 (2023).

22. Ioannidis, N. M. et al. REVEL: An Ensemble Method for Predicting the Pathogenicity of Rare Missense Variants. Am. J. Hum. Genet. 99, 877–885 (2016).

23. Sundaram, L. et al. Predicting the clinical impact of human mutation with deep neural networks. Nat. Genet. 50, 1161–1170 (2018).

24. Jaganathan, K. et al. Predicting Splicing from Primary Sequence with Deep Learning. Cell 176, 535–548.e24 (2019).

25. Tate, J. G. et al. COSMIC: the Catalogue Of Somatic Mutations In Cancer. Nucleic Acids Res. 47, D941–D947 (2019).

26. Namba, S. et al. Common Germline Risk Variants Impact Somatic Alterations and Clinical Features across Cancers. Cancer Res. 83, 20–27 (2023).

27. Kachuri, L. et al. Pan-cancer analysis demonstrates that integrating polygenic risk scores with modifiable risk factors improves risk prediction. Nat. Commun. 11, 6084 (2020).

28. Kim, E. S. et al. Potential utility of risk stratification for multicancer screening with liquid biopsy tests. Npj Precis. Oncol. 7, 39 (2023).

29. Hu, J., Ye, Y., Zhou, G. & Zhao, H. Using clinical and genetic risk factors for risk prediction of 8 cancers in the UK Biobank. JNCI Cancer Spectr. 8, pkae008 (2024).

30. Hu, C. et al. A Population-Based Study of Genes Previously Implicated in Breast Cancer. N. Engl. J. Med. 384, 440–451 (2021).

31. Zhou, W. et al. SAIGE-GENE+ improves the efficiency and accuracy of set-based rare variant association tests. Nat. Genet. 54, 1466–1469 (2022).

32. Karczewski, K. J. et al. Systematic single-variant and gene-based association testing of thousands of phenotypes in 394,841 UK Biobank exomes. Cell Genomics 2, 100168 (2022).

33. Engel, C. et al. BRAT1–related disorders: phenotypic spectrum and phenotype-genotype correlations from 97 patients. Eur. J. Hum. Genet. 31, 1023–1031 (2023).

34. So, E. Y. & Ouchi, T. The Potential Role of BRCA1-Associated ATM Activator-1 (BRAT1) in Regulation of mTOR. J. Cancer Biol. Res. 1, 1001 (2013).

35. Jurgens, S. J. et al. Adjusting for common variant polygenic scores improves yield in rare variant association analyses. Nat. Genet. 55, 544–548 (2023).

36. Wang, C. et al. Genome-wide analysis of runs of homozygosity identifies new susceptibility regions of lung cancer in Han Chinese. J. Biomed. Res. 27, 208–214 (2013).

37. Thomsen, H. et al. Runs of homozygosity and inbreeding in thyroid cancer. BMC Cancer 16, 227 (2016).

38. Lao, Y. et al. Association Between Alcohol Consumption and Risk of Bladder Cancer: A Dose-Response Meta-Analysis of Prospective Cohort Studies. Front. Oncol. 11, 696676 (2021).

39. SEER*Explorer Application. < https://seer.cancer.gov/statistics-network/explorer/application.html?site=72&data_type=5&graph_>type=11&compareBy=sex&chk_sex_1=1&series=9&&age_range=1&advopt_precision=2#resultsRegion1.

40. Hemminki, K., Sundquist, J. & Bermejo, J. L. How common is familial cancer? Ann. Oncol. 19, 163–167 (2008).

41. Lee, S. H., Wray, N. R., Goddard, M. E. & Visscher, P. M. Estimating missing heritability for disease from genome-wide association studies. Am. J. Hum. Genet. 88, 294–305 (2011).

42. Wilcox, N. et al. Exome sequencing identifies breast cancer susceptibility genes and defines the contribution of coding variants to breast cancer risk. Nat. Genet. 55, 1435–1439 (2023).

43. King, M.-C., Marks, J. H. & Mandell, J. B. Breast and Ovarian Cancer Risks Due to Inherited Mutations in BRCA1 and BRCA2. Science 302, 643–646 (2003).

44. McLaren, W. et al. The Ensembl Variant Effect Predictor. Genome Biol. 17, 122 (2016).

45. Frankish, A. et al. GENCODE reference annotation for the human and mouse genomes. Nucleic Acids Res. 47, D766–D773 (2019).

46. Jin, Y., Schaffer, A. A., Feolo, M., Holmes, J. B. & Kattman, B. L. GRAF-pop: A Fast Distance-Based Method To Infer Subject Ancestry from Multiple Genotype Datasets Without Principal Components Analysis. G3 9, 2447–2461 (2019).

47. Durbin, R. M. et al. A map of human genome variation from population-scale sequencing. Nature 467, 1061–1073 (2010).

48. Schneider, K., Zelley, K., Nichols, K. E., Levine, S. & Garber, J. Li-Fraumeni Syndrome.

49. Cunniff, C., Bassetti, J. A. & Ellis, N. A. Bloom’s Syndrome: Clinical Spectrum, Molecular Pathogenesis, and Cancer Predisposition. Mol. Syndromol. 8, 4–23 (2017).

50. Lauper, J. M., Krause, A., Vaughan, T. L. & Monnat, R. J. Spectrum and Risk of Neoplasia in Werner Syndrome: A Systematic Review. PLoS ONE 8, e59709 (2013).

51. Sasani, T. A. et al. Large, three-generation human families reveal post-zygotic mosaicism and variability in germline mutation accumulation. eLife 8, e46922 (2019).

52. Centre d’Etude du polymorphisme humain (CEPH): Collaborative genetic mapping of the human genome. Genomics 6, 575–577 (1990).

53. Han, S. Investigating the landscape of rare germline variants in cancer – from cancer risk to treatment response. https://nrs.harvard.edu/URN-3:HUL.INSTREPOS:37375611 (2023).

54. Bild, D. E. et al. Multi-Ethnic Study of Atherosclerosis: Objectives and Design. Am. J. Epidemiol. 156, 871–881 (2002).

55. Taliun, D. et al. Sequencing of 53,831 diverse genomes from the NHLBI TOPMed Program. Nature 590, 290–299 (2021).

56. Poplin, R. et al. Scaling accurate genetic variant discovery to tens of thousands of samples. 201178 Preprint at 10.1101/201178 (2018).

57. Lu, W. et al. CHARR efficiently estimates contamination from DNA sequencing data. Am. J. Hum. Genet. 110, 2068–2076 (2023).

58. Chen, S. et al. A genomic mutational constraint map using variation in 76,156 human genomes. Nature 625, 92–100 (2024).

59. Karczewski, K. J. et al. The mutational constraint spectrum quantified from variation in 141,456 humans. Nature 581, 434–443 (2020).

60. A global reference for human genetic variation. Nature 526, 68–74 (2015).

61. Mills, R. E. et al. An initial map of insertion and deletion (INDEL) variation in the human genome. Genome Res. 16, 1182–1190 (2006).

62. Hail Team. Hail.

63. Danecek, P. et al. The variant call format and VCFtools. Bioinformatics 27, 2156–2158 (2011).

64. Purcell, S. et al. PLINK: A Tool Set for Whole-Genome Association and Population-Based Linkage Analyses. Am. J. Hum. Genet. 81, 559–575 (2007).

65. Gillani, R. et al. Rare germline structural variants increase risk for pediatric solid tumors. Science 387, eadq0071 (2025).

66. Liu, X., Jian, X. & Boerwinkle, E. dbNSFP: A Lightweight Database of Human Nonsynonymous SNPs and Their Functional Predictions. Hum. Mutat. 32, 894–899 (2011).

67. Khera, A. V. et al. Genome-wide polygenic scores for common diseases identify individuals with risk equivalent to monogenic mutations. Nat. Genet. 50, 1219–1224 (2018).

68. Collins, R. L. et al. A cross-disorder dosage sensitivity map of the human genome. Cell 185, 3041–3055.e25 (2022).

69. Narasimhan, V. et al. BCFtools/RoH: a hidden Markov model approach for detecting autozygosity from next-generation sequencing data. Bioinformatics 32, 1749–1751 (2016).

70. Loh, P.-R. et al. Reference-based phasing using the Haplotype Reference Consortium panel. Nat. Genet. 48, 1443–1448 (2016).

71. Quinlan, A. R. & Hall, I. M. BEDTools: a flexible suite of utilities for comparing genomic features. Bioinformatics 26, 841–842 (2010).

72. Miga, K. H. et al. Centromere reference models for human chromosomes X and Y satellite arrays. Genome Res. 24, 697–707 (2014).

73. Sondka, Z. et al. The COSMIC Cancer Gene Census: describing genetic dysfunction across all human cancers. Nat. Rev. Cancer 18, 696–705 (2018).

74. Oriola, J. et al. Novel truncating germline variant reinforces TINF2 as a susceptibility gene for familial non-medullary thyroid cancer. J. Med. Genet 61, 939–942 (2024).

