## Supplementary Figures 1-11 for "Diverse mediators of cancer predisposition uncovered by germline whole genome sequencing of unexplained familial cancers"

### Supplementary Information for Fields *et al.*

### SUPPLEMENTARY TABLES

1. List of 148 CPGs considered in this study
2. Mapping of EHR cancer diagnosis codes to cancer type categories evaluated in this study
3. Mapping of 79 CPGs to COSMIC cancer phenotypes
4. Cancer Type-PRS matrix for p-values
5. Cancer Type-PRS matrix for OR
6. Exome-wide rare variant gene association statistics for 15 cancer types (available upon request)
7. Genome-wide Runs of Homozygosity statistics for bladder, kidney, and thyroid cancer (available upon request)

### SUPPLEMENTARY FIGURES

**Supplementary Figure 1. Environmental risk factors in our cohort.**

**Supplementary Figure 2. Quality control metrics for germline SNVs and indels as a function of true positive probability threshold calculated from our random forest filtering model.**

**Supplementary Figure 3. Germline variant summary metrics.**

**Supplementary Figure 4. Inference of genetic ancestry and sex.**

**Supplementary Figure 5. Variant counts stratified by ancestry.**

**Supplementary Figure 6. Distribution of polygenic risk across unexplained familial cancer cases, stratified by concordant family history.**

**Supplementary Figure 7. Distribution of polygenic risk across unexplained familial cancer cases with a family history of the PRS-matched cancer type.**

**Supplementary Figure 8. Definitions for rare genic and splicing SNVs/indels using ClinVar annotations and *in silico* variant effect predictors.**

**Supplementary Figure 9. Quantile-quantile plots of exome-wide predisposition gene discovery analyses with SAIGE-GENE+.**

**Supplementary Figure 10. Genome-wide manhattan plots of RoH association statistics for two cancer types with at least one significant RoH association.**

**Supplementary Figure 11. The contribution of germline genetic factors including novel risk-associated loci beyond canonical PGVs to familial cancer liability.**

**REFERENCES**

1. Durbin, R. M. et al. A map of human genome variation from population-scale sequencing. Nature 467, 1061–1073 (2010).
2. Jin, Y., Schaffer, A. A., Feolo, M., Holmes, J. B. & Kattman, B. L. GRAF-pop: A Fast Distance-Based Method To Infer Subject Ancestry from Multiple Genotype Datasets Without Principal Components Analysis. G3 9, 2447–2461 (2019).
3. Collins, R. L. et al. A structural variation reference for medical and population genetics. Nature 581, 444–451 (2020).
4. Miyado, M. & Fukami, M. Losing maleness: Somatic Y chromosome loss at every stage of a man's life. FASEB BioAdvances 1, 350–352 (2019).

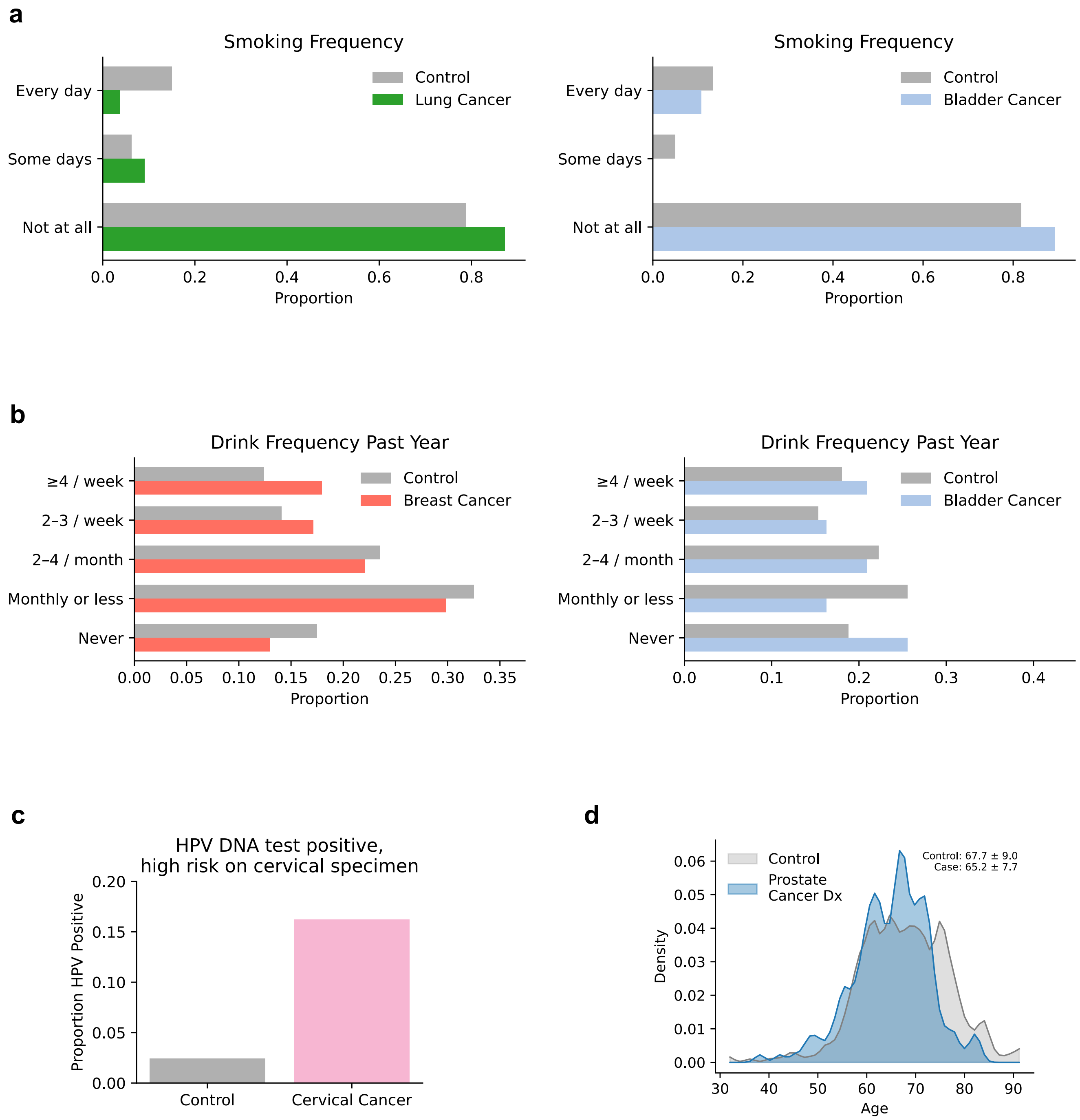


**Supplementary Figure 1. Environmental risk factors in our cohort.**

***(a)** Smoking distribution in individuals with lung and bladder cancers compared to matched controls. **(b)** Alcohol consumption rates in individuals with breast and bladder cancers compared to matched controls. **(c)** Proportion of individuals with a positive high-risk human papillomavirus (HPV) DNA test among cervical cancer cases and matched controls. **(d)** Age distribution of prostate cancers compared to matched controls. These plots display proportions after excluding individuals with missing or incomplete survey data. To comply with the All of Us Research Program's data and statistics dissemination policies, we do not report absolute counts for individual response categories.*

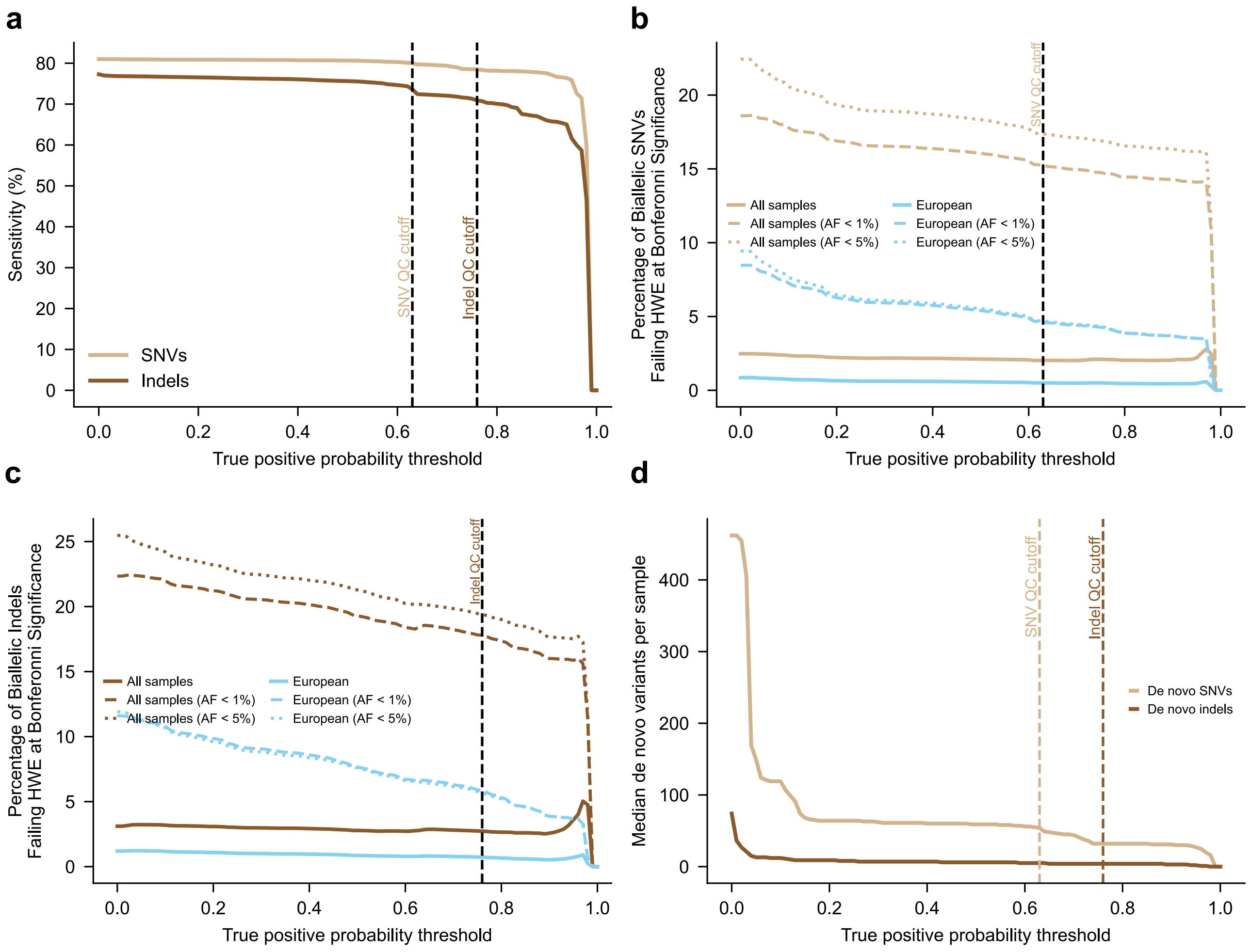


### Supplementary Figure 2. Quality control metrics for germline SNVs and indels as a function of true positive probability threshold calculated from our random forest filtering model.

***(a)*** *The percentage of 1000 Genomes Project SNVs and indels that were captured in our analysis*[1]. ***(b-c)*** *The percentage of biallelic* ***(b)*** *SNVs and* ***(c)*** *indels that significantly deviated from Hardy–Weinberg equilibrium (HWE), where Bonferroni-adjusted significance thresholds were recalculated for each combination of variant type, allele frequency, and ancestry.* ***(d)*** *Median rate of de novo SNVs and indels in 367 complete parent-child trios.*

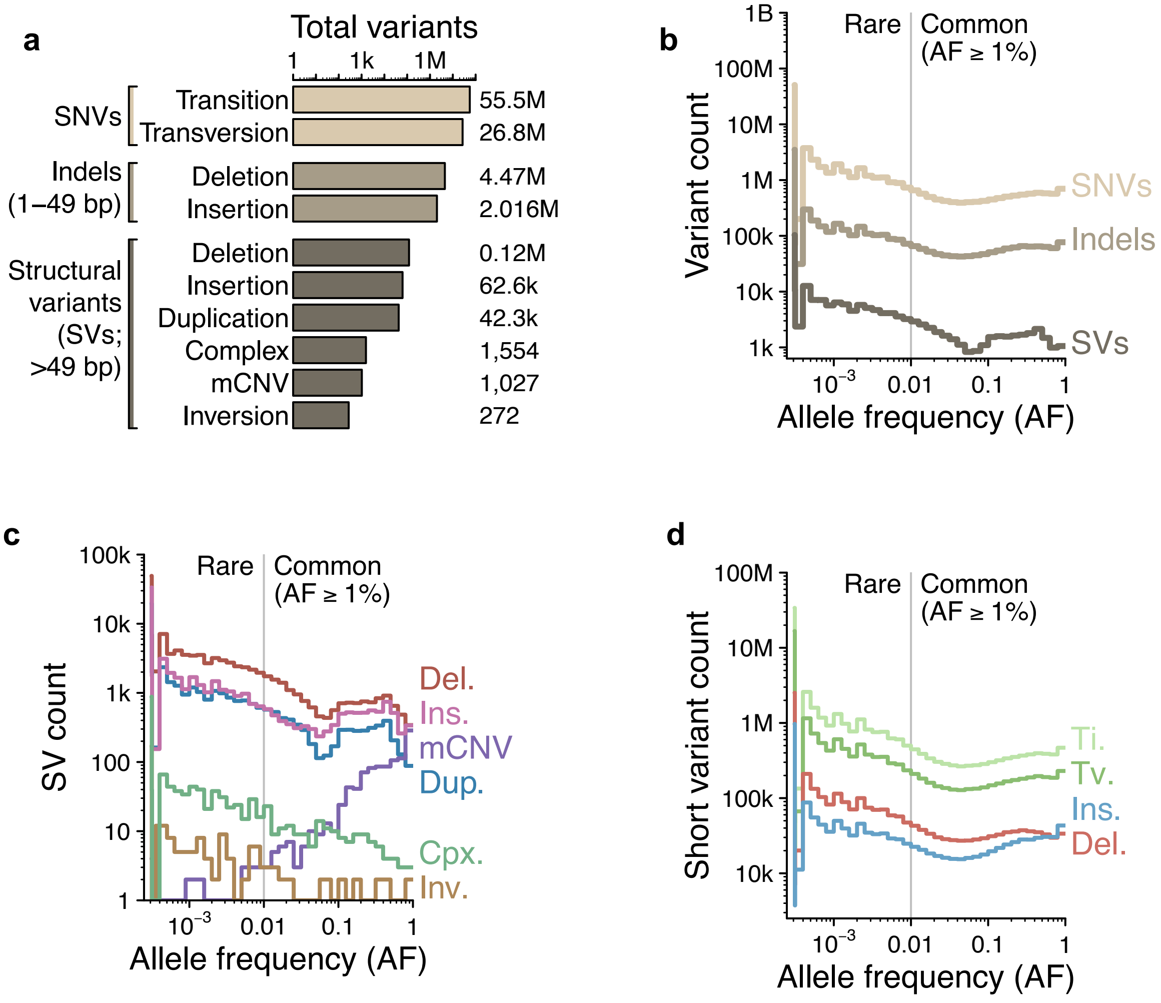


**Supplementary Figure 3. Germline variant summary metrics.**

*__(a)__ Genome-wide variant counts for all 2,695 individuals in our cohort who passed strict structural variant (SV) quality control filtering (99% of cohort). mCNV., multiallelic copy number variant. __(b)__ SNV, indel, and SV counts across variant allele frequencies within our cohort. __(c)__ The distribution of SV types across variant allele frequencies. __(d)__ The distribution of SNV/indel types across variant allele frequencies.*

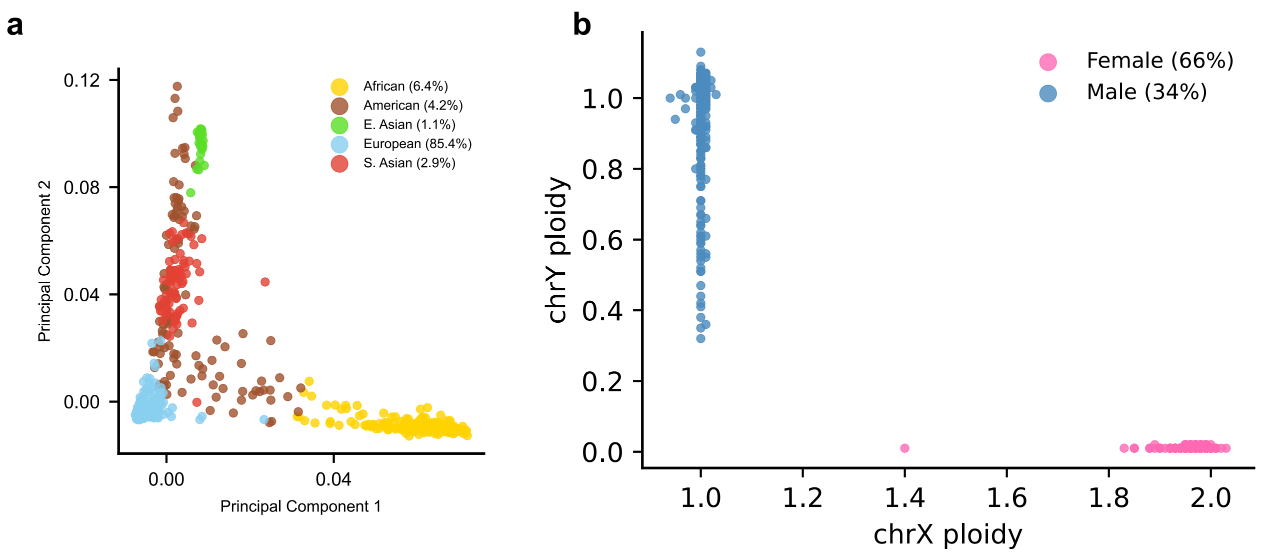


**Supplementary Figure 4. Inference of genetic ancestry and sex.**

**(a)** We performed principal components analysis (PCA) on common, well-genotyped, biallelic, LD-pruned, SNVs in our filtered germline variant callset to discern genetic ancestry. Genetic ancestry was predicted with GRAF-pop[2]. (**b**) Sex inference from GATK-SV for the 2,726 individuals in our cohort used for formal association testing[3]. Less than 20 cases in our cohort were found to have sex aneuploidies and were thus not included in this graph to abide by the All of Us Research Program's data dissemination and statistics policy. We observe a clear signature of age-related mosaic loss-of-Y in blood, which is a known phenomenon[4].

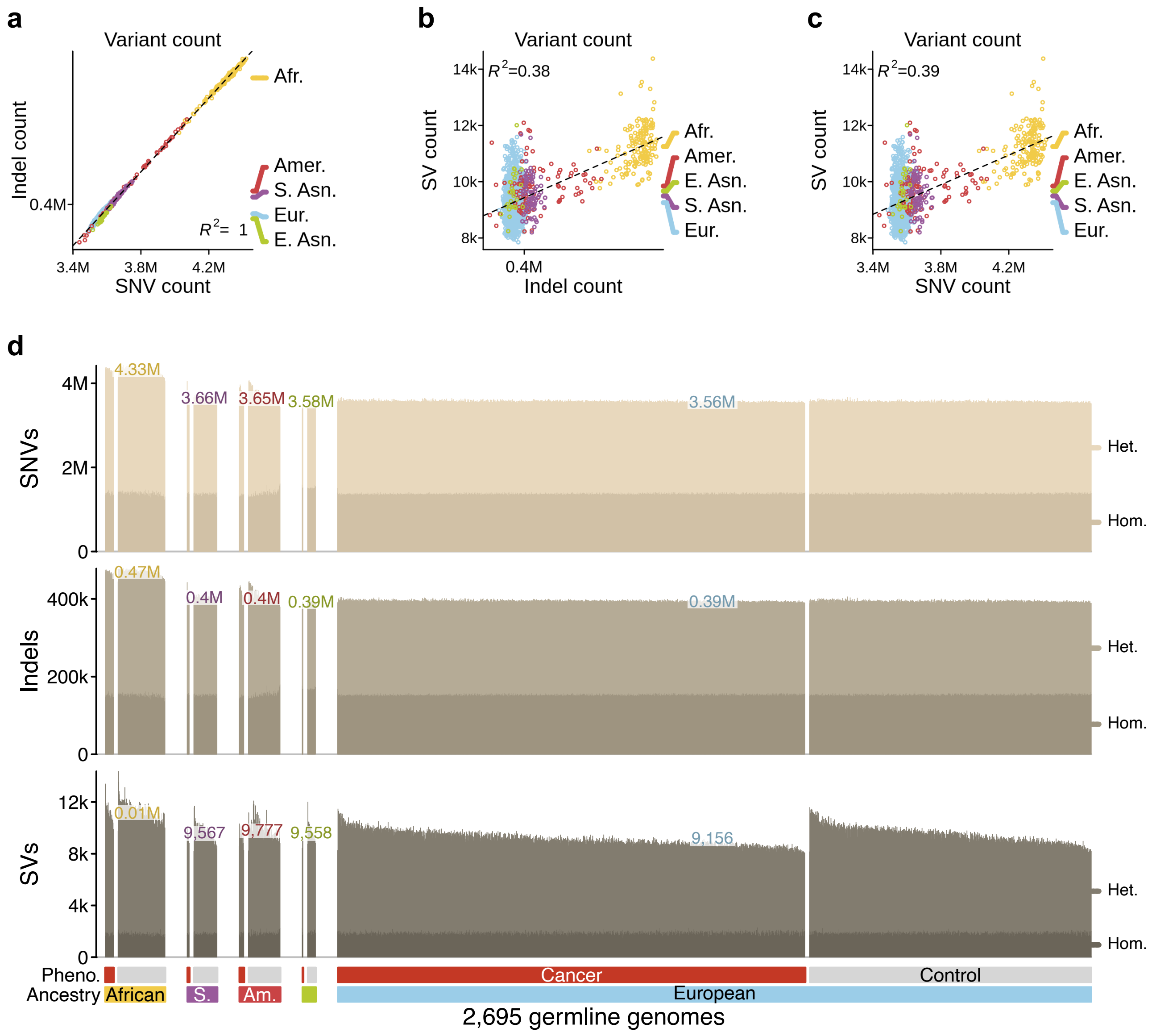


### Supplementary Figure 5. Variant counts stratified by ancestry.

***(a)*** *Comparison of SNV and indel counts per genome.* ***(b)*** *Comparison of indel and SV counts per genome.* ***(c)*** *Comparison of SNV and SV counts per genome.* ***(d)*** *Total count of SNVs, indels, and SVs per genome stratified by genetic ancestry and case-control status. Heterozygosity and homozygosity is indicated by bar saturation per the right axis.*

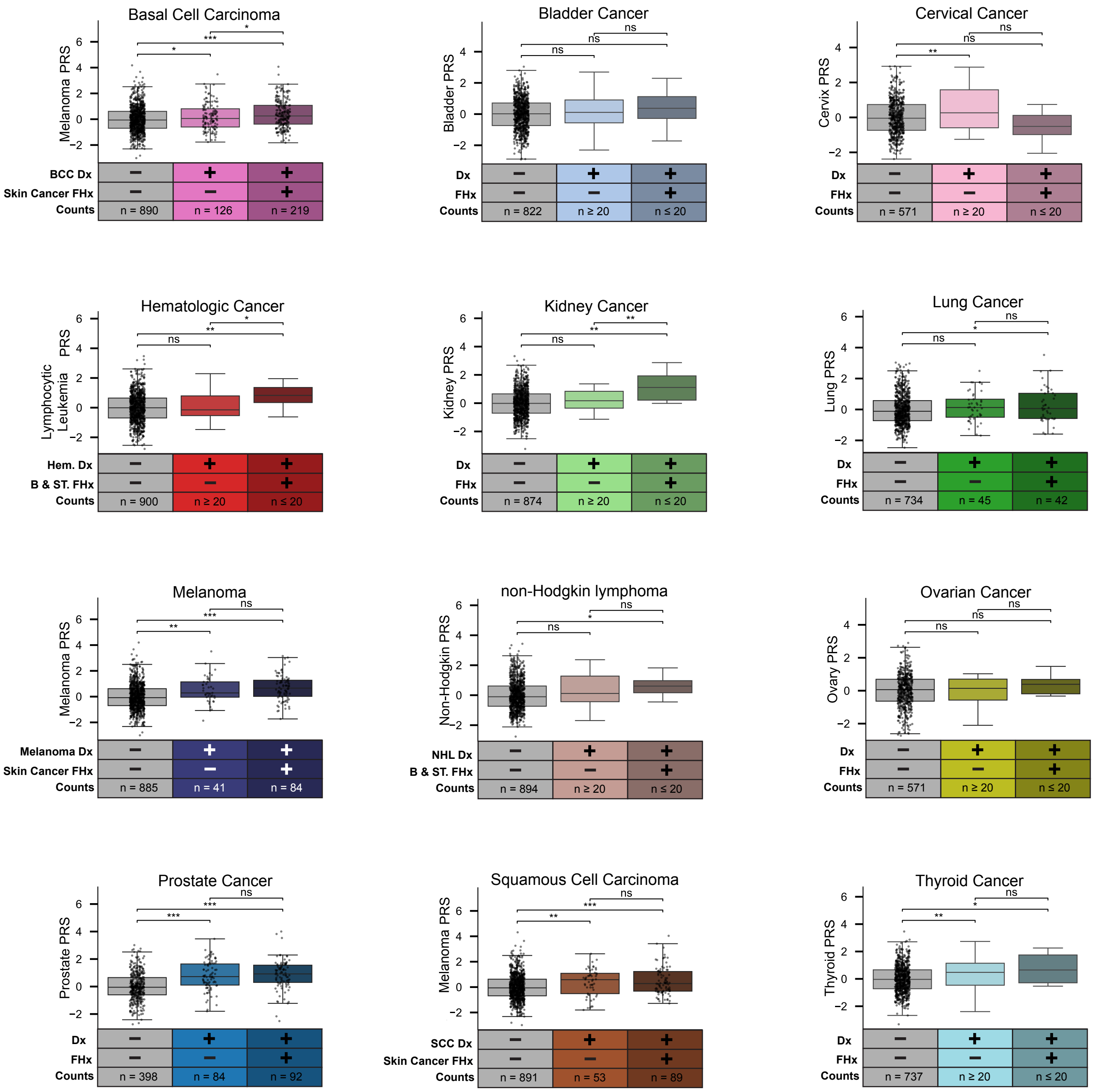


**Supplementary Figure 6. Distribution of polygenic risk across unexplained familial cancer cases, stratified by concordant family history.**

*Plots show the distribution of familial cancer cases across cancer types. Statistical significance is indicated as follows: P < 0.05 (*), P < 0.01 (**), P < 1 × 10-5 (***), and not significant (ns). Within each cancer type, cases are further subdivided based on family history of the PRS-matched cancer. Family history refers to a first-degree relative with the corresponding cancer.*

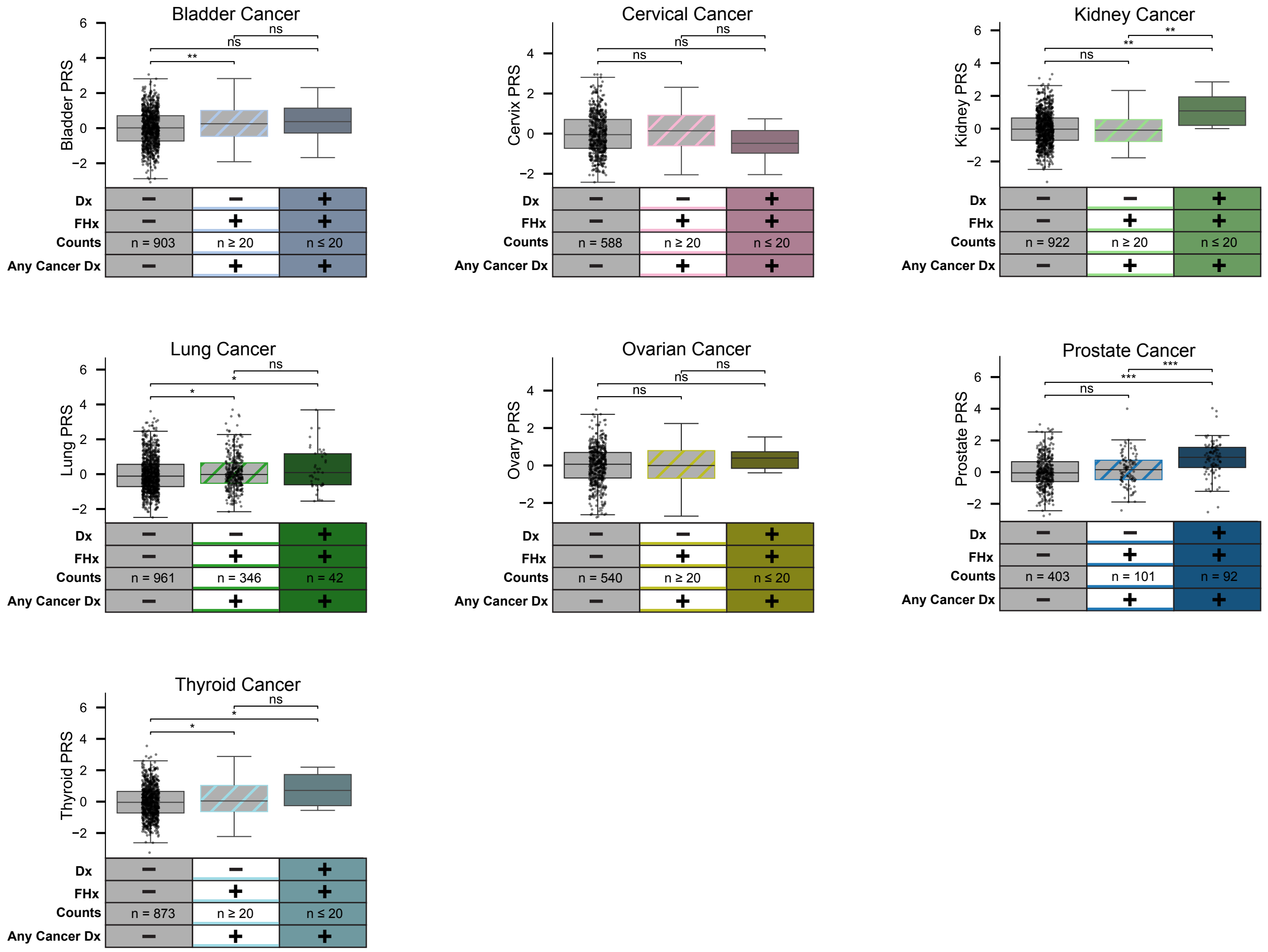


### Supplementary Figure 7. Distribution of polygenic risk across unexplained familial cancer cases with a family history of the PRS-matched cancer type.

*Plots show the distribution of familial cancer cases across cancer types. Statistical significance is indicated as follows: P < 0.05 (*), P < 0.01 (**), P < 1 × 10-5 (***), and not significant (ns). Within each cancer type, cases are defined as those with any cancer diagnosis and a family history of the PRS-matched cancer. We further subdivided based on specific cancer type diagnosis. Family history refers to a first-degree relative with the corresponding cancer.*

| Name | Variant Inclusion Criteria | Label | Median Count per Genome (AF ≤ 0.01) |
|---|---|---|---|
| Tier 1 Variant | VEP HIGH Impact & (LOFTEE HC OR ClinVar P/LP)* | High Confidence LOF | 14 |
| Tier 2 Variant | LOFTEE Low Confidence OR SpliceAI ≥ 0.5 OR (SpliceAI ≥ 0.2 & ClinVar P/LP)* | pLOF / Splice-Disruptive | 14 |
| Tier 3 Variant | 2 of (PrimateAI, AlphaMissense, and REVEL ≥ 0.75) OR ClinVar P/LP Missense Variant* | Predicted Damaging Missense | 13 |
| Tier 4 Variant** | 1 of (PrimateAI, AlphaMissense, and REVEL ≥ 0.5) OR ClinVar P/LP Missense Variant* | Weakly Damaging Missense | 58 |
| Tier 5 Variant | 0 of (PrimateAI, AlphaMissense, and REVEL ≥ 0.5) | Neutral Missense | 207 |
| Tier 6 Variant | Synonymous | Synonymous | 190 |

* Removed Variants with ClinVar Benign or Likely Benign annotations
** Tier 4 is a superset of Tier 3 Variants

**Supplementary Figure 8. Definitions for rare genic and splicing SNVs/indels using ClinVar annotations and in silico variant effect predictors.**

*We enumerated 6 heuristic-based tiers to group rare variants based on predicted severity of consequence on gene function or carrier phenotype. On the right, we denoted the median count per genome of variants for AF < 1%. This tier system was used for our rare-variant gene-burden analysis using SAIGE-GENE+.*

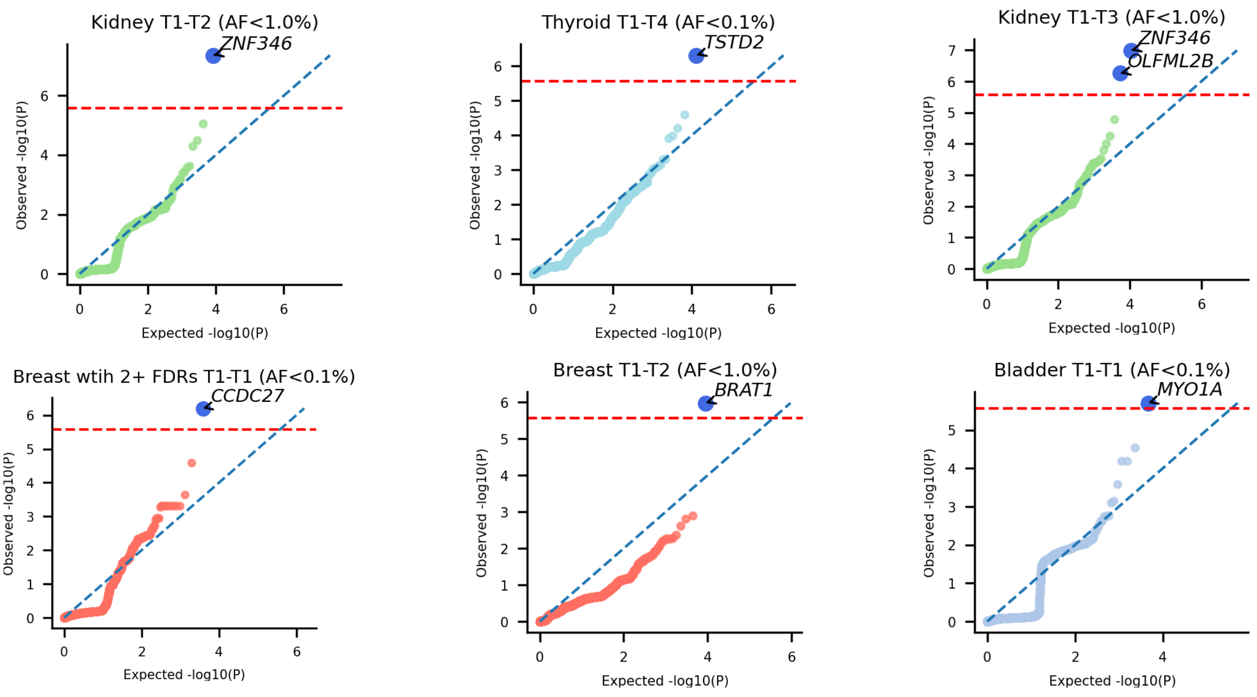


**Supplementary Figure 9. Quantile-quantile plots of exome-wide predisposition gene discovery analyses with SAIGE-GENE+.**

*Genes that reached exome-wide thresholds with GeneBass evidence are colored in blue and labeled. The dashed red line denotes exome-wide significance (P < 2.69 x $10^{-6}$).*

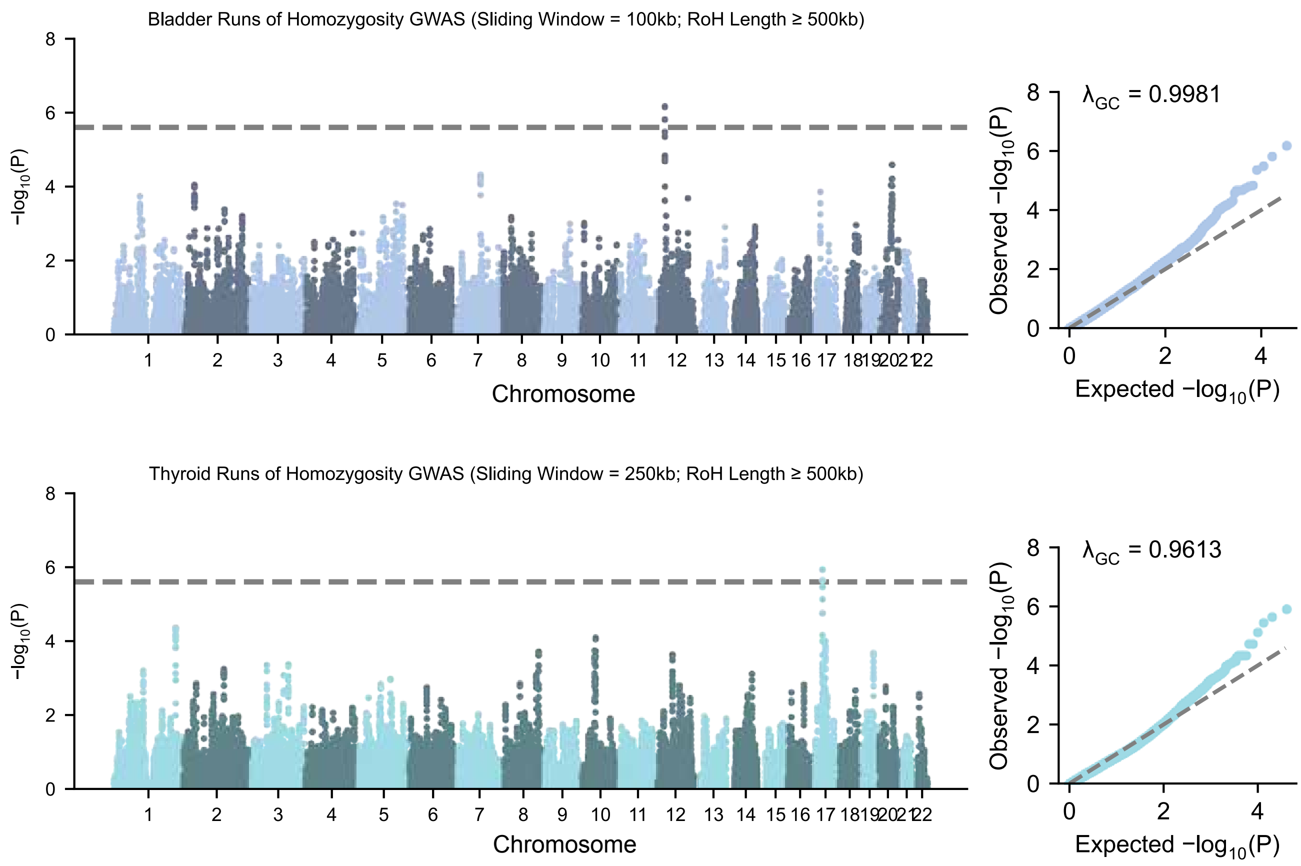


**Supplementary Figure 10. Genome-wide manhattan plots of RoH association statistics for two cancer types with at least one significant RoH association.**

*Results from genome-wide RoH association testing for bladder cancer and thyroid cancer showing genome-wide significant loci at chr12:22,150,001–22,250,000 and chr17:30,150,001–30,400,000, respectively.*

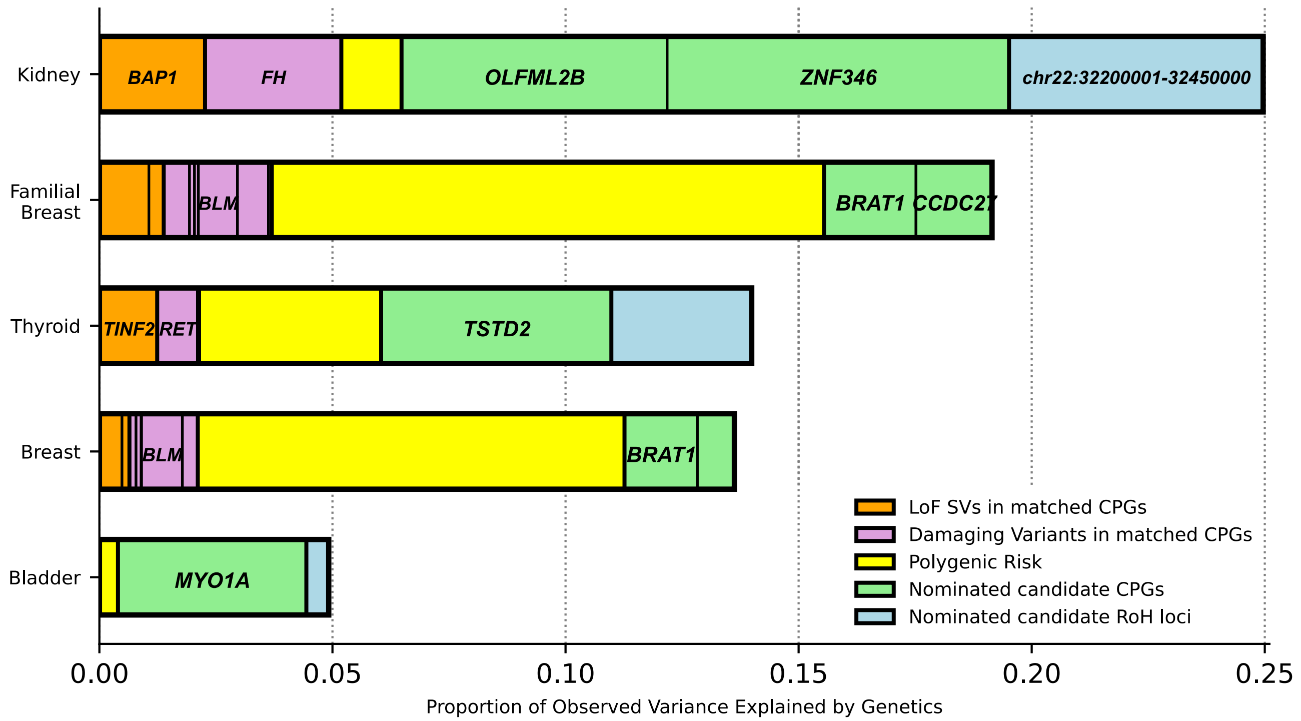


**Supplementary Figure 11. The contribution of germline genetic factors including novel risk-associated loci beyond canonical PGVs to familial cancer liability.**

*Proportion of observed phenotypic variance explained across 4 cancer types in our study when incorporating underappreciated classes of variation involving known cancer risk loci: LoF SVs in five selected genes; rare SNVs/indels in cancer type-matched CPGs; PRS; nominated CPGs; and nominated RoH Loci.*
